# Intramuscular SARS-CoV-2 vaccines elicit varying degrees of plasma and salivary antibody responses as compared to natural infection

**DOI:** 10.1101/2021.08.22.21262168

**Authors:** George Ronald Nahass, Rachel E. Salomon-Shulman, Grace Blacker, Kazim Haider, Rich Brotherton, Kristine Teague, Ying Ying Yiu, Rachel E. Brewer, Sarah Danielle Galloway, Paige Hansen, Gabriel Marquez-Arreguin, Salma Sheikh-Mohamed, Gary Y.C. Chao, Baweleta Isho, Evan Do, Iris Chang, Theo Snow, Alexandra S. Lee, STANFORD COVID-19 BIOBANK, Monali Manohar, Samuel Yang, Andra L. Blomkalns, Angela J Rogers, Allison McGeer, Anne-Claude Gingras, Sharon Straus, Phillip Grant, Kari C. Nadeau, Catherine A. Blish, Jennifer L. Gommerman, Erin C. Sanders, Irving L. Weissman, Michal Caspi Tal

## Abstract

Vaccination induced antibody and T-cell immune responses are important for systemic protection from COVID-19. Because SARS-CoV-2 infects and is transmitted by oral-pharyngeal mucosa, we wished to test mucosal antibodies elicited by natural infection or intramuscular vaccine injection. In a non-randomized observational study, we measured antibodies against the SARS-CoV-2 RBD in plasma and saliva from convalescent or vaccinated individuals and tested their neutralizing potential using a replication competent rVSV-eGFP-SARS-CoV-2. We found IgG and IgA anti-RBD antibodies as well as neutralizing activity in convalescent plasma and saliva. Two doses of mRNA vaccination (BNT162b2 or mRNA-1273) induced high levels of IgG anti-RBD in saliva, a subset of whom also had IgA, and significant neutralizing activity. We detected anti-RBD IgG and IgA with significant neutralizing potential in the plasma of single dose Ad26.COV2.S vaccinated individuals, and we detected slight amounts of anti-RBD antibodies in matched saliva. The role of salivary antibodies in protection against SARS-CoV-2 infection is unknown and merits further investigation. This study was not designed to, nor did it study the full kinetics of the antibody response or protection from infection, nor did it address variants of SARS-CoV-2.

## INTRODUCTION

The rapid and successful development of multiple highly effective and safe vaccines to fight the pandemic caused by Severe Acute Respiratory Syndrome Coronavirus 2 (SARS-CoV-2) is predicated on years of existing work and remarkable (1–3). Three vaccines are currently granted Emergency Use Authorization (EUA) by the United States (US) Food and Drug Administration (FDA) and are being administered to the American public: BNT162b2 from Pfizer/BioNTech (*4*), mRNA-1273 from ModernaTX, Inc (*5*), and Ad26.COV2.S from Johnson & Johnson/Janssen Pharmaceuticals (J&J) (*6*). BNT162b2 and mRNA-1273 are two dose messenger ribonucleic acid (mRNA) vaccines, while Ad26.COV2.S utilizes a human adenovirus vector in a single dose. All three vaccines stimulate antibodies to the SARS-CoV-2 spike “S” protein, which are capable of virus neutralization and other functions. Additionally, in clinical trials, all three vaccines induce robust T-cell immunity and significantly reduced the rate of symptomatic coronavirus disease 2019 (COVID-19) (4–6).

SARS-CoV-2 is an aerosol transmissible virus, which upon inhalation into the mucosa of the nasopharynx and oral cavity, infects locally and disseminates systemically, progressing into pulmonary and multi-organ infection (*7–9*). The human nasal and oral epithelium have been shown to highly express SARS-CoV-2 entry factors *in vitro* (*10–12*) and studies using animal models have shown the ability of SARS-CoV-2 to infect and replicate in the olfactory epithelium of the nasal turbinates in mice (*13, 14*), hamsters (*15–18*) and rhesus macaques (*7*). Consequently, both infection and transmission of infection occur via the oral-pharyngeal mucosa.

The objective of this study was to evaluate whether intramuscular injection of vaccination could induce a mucosal antibody response. To assess this question, we evaluated the antibodies present in saliva or plasma of study participants after infection or vaccination against SARS-CoV-2. This study was observational on samples provided by collaborators; our part of the study was not prospective, and therefore was limited by sample availability.

The route of vaccination influences the site of immune responses. In general, the intramuscular route induces systemic immunity in circulation, while intranasal or oral infection or vaccination induces mucosal antibodies that can enter the fluids bathing the mucosa (*3, 19*). Compared with a nasal swab, saliva is a unique specimen to evaluate, as it is readily accessible, contains secreted mucosal antibodies, and offers a glimpse into circulating antibodies, attributed to vascular leakage from the gingival crevicular epithelium (*20, 21*). In saliva, the two major antibody classes are Immunoglobulin A (IgA) and Immunoglobulin G (IgG). Plasma cells (PCs) or their precursors from the draining lymphoid organs of infected or vaccinated mucosal surfaces migrate to the local mucosa and produce secretory IgA (SIgA) with a joining “J” chain to make IgA dimers, and then pairs with a secretory component (SC) to help cross the epithelial cell cytoplasm. This dimeric SIgA may play a role in protecting the mucosal surfaces (20–22). A small amount of IgG is also produced locally by the gingival, glandular, and tonsillar PCs (*20*). Antibodies in the serum and saliva can be neutralizing (able to bind to a pathogen and prevent it from infecting host cells) or non-neutralizing (*20, 21, 23*).

T-cells are also known to play an important role in viral infection with both effector cells such as CD8+ T-cells and memory T-cells, as well as helping or regulating B-cells and their progeny plasma cells in generating antibody responses (*24*). As our study was restricted to the role of antibodies, T-cells were not evaluated in this study, though they may play a role that is especially important in protection against severe disease resulting from new variants.

Symptomatic SARS-CoV-2 infection typically induces a robust systemic antibody response corresponding with protection from disease (*25–28*). In SARS-CoV-2 infection, IgA can be important in the neutralizing antibody (NAb) response (*29*). Early in the pandemic, researchers in China found that SARS-CoV-2 was detected in the saliva of 91.7% of hospitalized patients with COVID-19 from which live virus was able to be grown in culture (*30*). This finding is consistent with the early detection and high viral loads of another viral infection, SARS-CoV-1, in the saliva of hospitalized patients, even before the development of lung lesions (*31*). Wyllie et al demonstrated that saliva has been an appropriate body fluid to screen for SARS-CoV-2 infection and equivalent to nasopharyngeal swab testing (*32*). Direct correlation between paired saliva and blood samples from patients with COVID-19 also demonstrates how the systemic immune response can be monitored through saliva (*33*).

Recent studies have evaluated serum and mucosal tissues including saliva and nasal swabs for cellular and antibody immunity to COVID-19 and vaccination and shown that a multifaceted immune response is important for protection (*26, 34–39*). Further, there is evidence that NAbs are an important correlate of protection in the immune response to SARS-CoV-2 (*26, 38–42*).

Given our current understanding of mucosal immunity and its significance to the COVID-19 pandemic, the question remains whether an intramuscular vaccination can induce an immune response that prevents infection and transmission in the local mucosa. In an observational study we examined antibody responses from matched plasma and saliva samples (Sup. Figure 1) in a longitudinal cohort of participants enrolled in the J&J Phase 3 ENSEMBLE trial of the Ad26.COV2.S vaccine (Sup. Table 1) and in individuals who received BNT162b2 or mRNA-1273 vaccines (Sup. Table 2). These observational studies were not prospectively designed and were carried out as a pilot study to compare salivary and plasma antibodies to the receptor binding domain (RBD) of SARS-CoV-2 infection or vaccination. We could therefore not provide timing or subgrouping of cohorts to analyze the responses thoroughly, but the potential for learning how much and which kinds of antibodies to the viral RBD are present in the oral cavity as compared to the plasma of vaccinated or infected individuals were of sufficient importance to warrant this pilot study. The results warrant more thorough prospectively designed and randomized studies to expand upon this research.

**Fig. 1:**
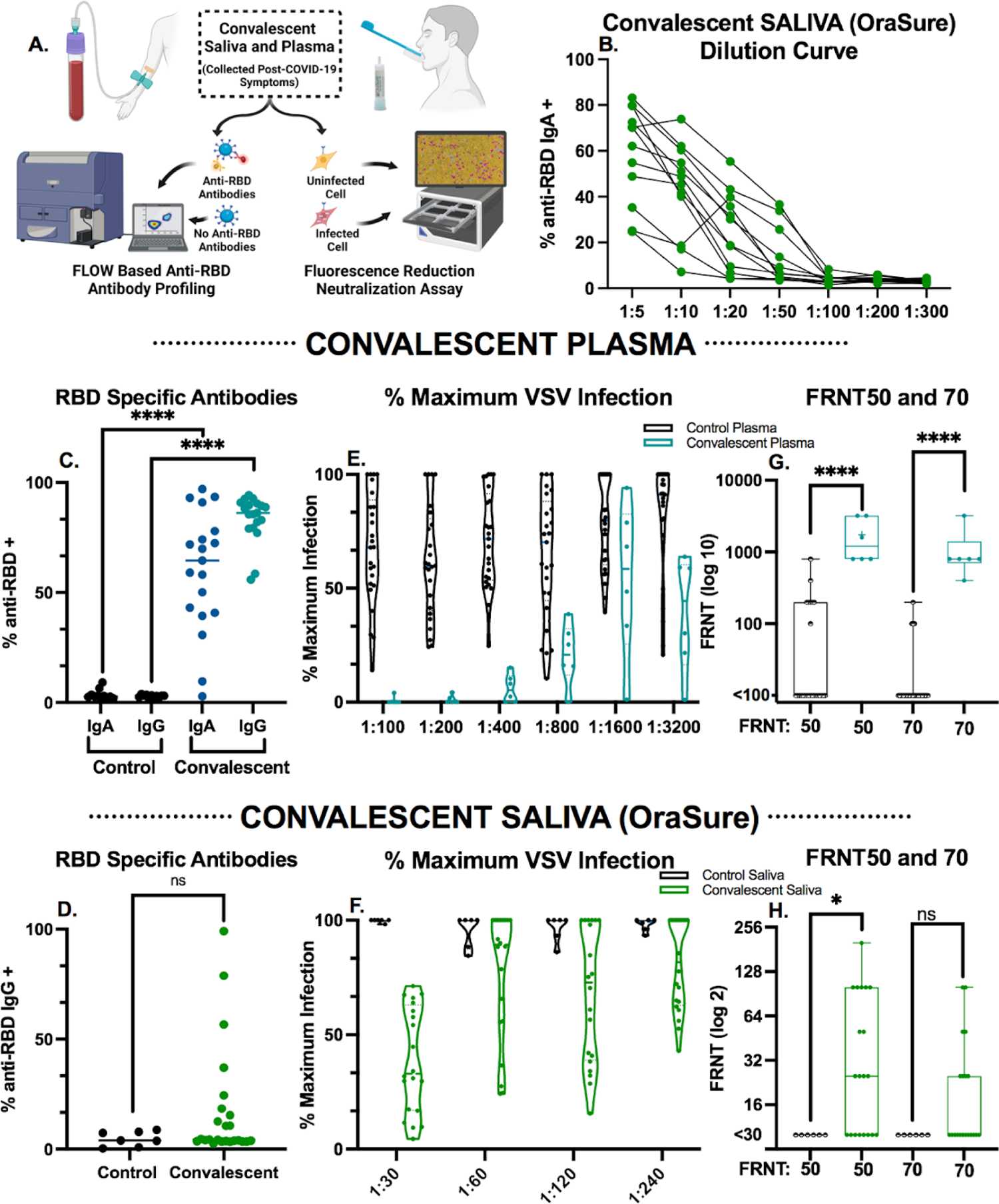
Convalescent saliva and plasma both have detectable RBD-specific antibodies and neutralization activity. **A:** Schematic of the sample collection and downstream analysis. **B:** Serial dilution of saliva measured for RBD-specific IgA antibodies by flow cytometry. Each symbol indicates a particular sample, for comparison across dilutions. **C:** Plasma from pre-pandemic controls compared to COVID-19 convalescent plasma utilized to set thresholds in all flow cytometry experiments diluted 1:900 and assessed for RBD-specific antibodies as indicated by flow cytometry Mann-Whitney U tests were used for comparisons (*\*\*\*\*p* ≤.0001). **D:** Convalescent saliva (n=20) compared to known negative controls (n=7) utilized to set thresholds in all flow cytometry experiments diluted 1:10 and assessed for RBD-specific antibodies as indicated by flow cytometry Mann-Whitney U tests were used for comparisons (ns *p*>.05). **E-F:** Neutralization activity from convalescent E) plasma (n=6) or F) saliva (n=20) from patients who recovered from COVID-19 represented as percent of maximum of the triplicate rVSV-eGFP-SARS-CoV-2 control conditions. Each dot represents the fluorescent signal from rVSV-eGFP-SARS-CoV-2 infection from an individual patient in the assay. Any values over 100% of maximum infection were deemed as saturated and as such a threshold of 100% was set. Plasma (E) and saliva (F) samples were analyzed at two step dilutions ranging from 1:100-1:3200 and 1:30-1:240 respectively. **G-H:** Fluorescence reduction of neutralization titer (FRNT) of rVSV-eGFP-SARS-CoV-2 infection of HEK293-hACE2-mCherry expressing cells from convalescent G) Plasma (n=6) or H) saliva (n=20) samples compared to control saliva or plasma respectively. For each step in the serial dilutions, fluorescence intensity of the sample as a % of the average of the fluorescent signal of triplicate rVSV-eGFP-SARS-CoV-2 only wells is indicated as % of maximum. FRNT50 and FRNT70 represent the minimum dilution that had 50% or 30%, respectively, of the average of the fluorescent signal of triplicate rVSV-eGFP-SARS-CoV-2 control wells. Box plots represent the median and interquartile range and whiskers extend to the maximum or minimum values. Mann-Whitney U tests were used for comparisons (\*\**p* ≤.01, \*\*\*\**p* ≤.0001).

## RESULTS

Overall, we studied the antibodies in plasma and/or saliva of participants after vaccination against SARS-CoV-2 with either Ad26.COV2.S, BNT162b2, or mRNA-1273 vaccines (Sup. Tables 1, 2, and 5, respectively) or after natural infection with SARS-CoV-2 (Sup. Tables 3 and 4, respectively). For this study, we compared four different methods for saliva collection to evaluate ease of sampling and antibody content. All devices compared require contact with mucosal tissues as well as saliva, except for passive drool. OraSure saliva collection devices were selected for longitudinal sampling as they recovered the highest number of antibody isotypes and subtypes (Sup. Figure 2) by flow cytometric analysis and contained a thermostable preservative that allowed for 21 days between collection and processing. Plasma samples collected in 2018 (demographic data was not provided for pre-pandemic samples) prior to the COVID-19 pandemic (n=11) and saliva samples (n=7) that were RT-PCR negative for SARS-CoV-2 with no known history of infection with SARS-CoV-2 (Sup. Table 6) herein referred to as “known negative,” were used as negative controls. Plasma and saliva samples collected from individuals recovered from acute SARS-CoV-2 infection (Sup. Tables 3 and 4) were evaluated for RBD-specific antibodies and Spike neutralizing activity (Figure 1A) and used as positive controls.

**Fig. 2:**
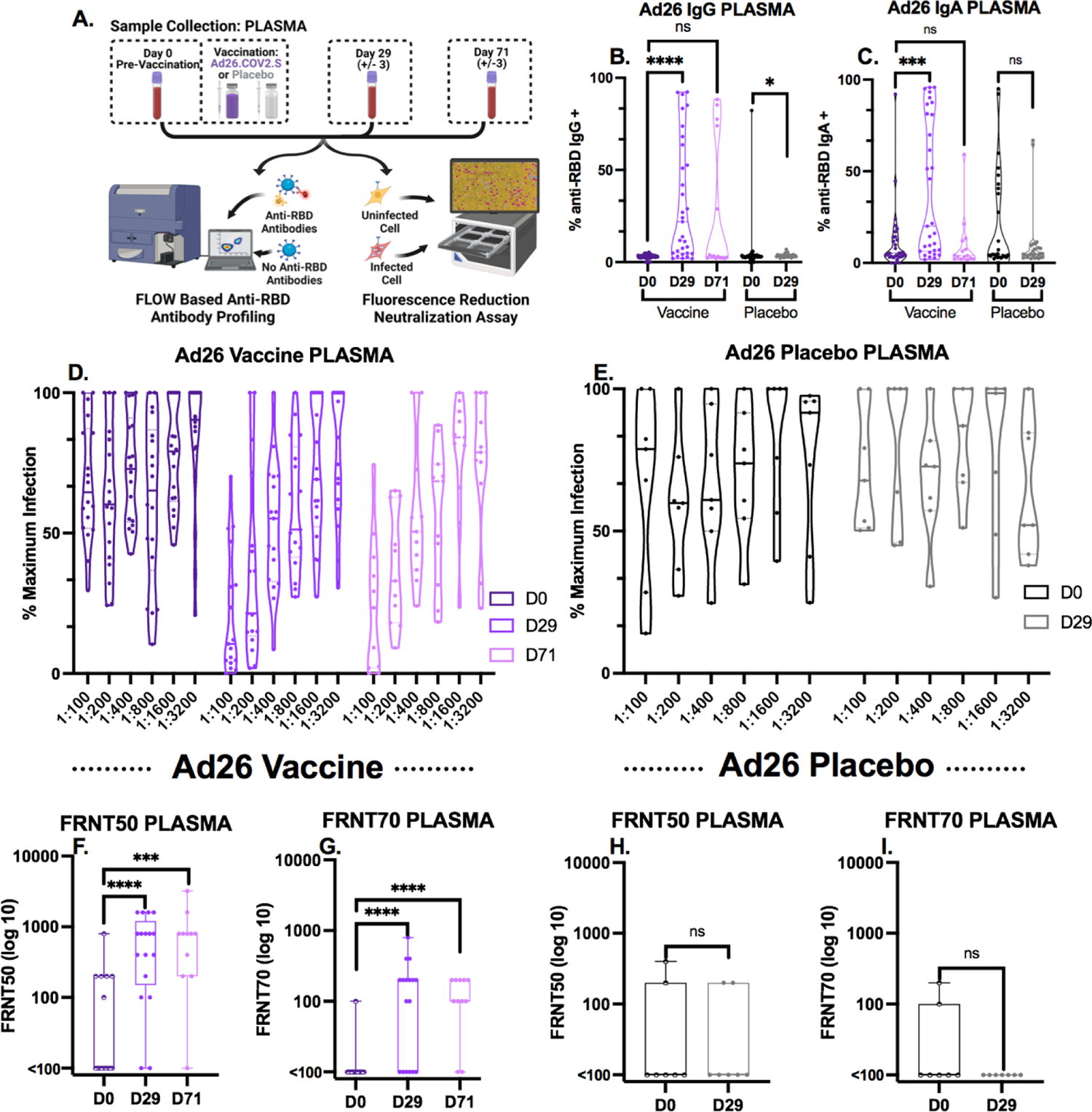
Ad26.COV2.S vaccinated individuals have high levels of RBD-specific antibodies and significantly increased neutralization activity in plasma. **A**: Schematic of plasma collection timeline and downstream analysis. **B-C:** Levels of RBD-specific B) IgG and C) IgA in the plasma of recipients of Ad26.COV2.S (n=31) or a placebo saline injection (n=27) measured by flow cytometry. Box plots represent the median and interquartile range and whiskers extend to the maximum or minimum values. Mann-Whitney U tests were used for comparisons (ns *p>*.05, \**p*≤ .05, \*\*\**p*≤.001, *\*\*\*\*p* ≤.0001). **D-E:** Neutralization activity of plasma from D) individuals vaccinated with Ad26.COV2.S (n=18 for D0 and D29, n=11 for D71) or E) a placebo saline injection (n=7) represented as percent of maximum of triplicate rVSV-eGFP-SARS-CoV-2 control conditions. Each dot represents the fluorescent signal from rVSV-eGFP-SARS-CoV-2 infection from an individual participant sample in the assay. Any values over 100% of maximum infection were deemed as saturated and as such a threshold of 100% was set. Plasma samples were analyzed at two step dilutions ranging from 1:100-1:3200. **F-I:** Fluorescence reduction of neutralization titer (FRNT) 50 and 70 of rVSV-eGFP-SARS-CoV-2 infection of HEK293-hACE2-mCherry expressing cells from plasma of F-G) individuals vaccinated with AD.26.COV2.S (n=18 for D0 and D29, n=11 for D71) or H-I) a placebo saline shot (n=7). See Figure 1 for details on analysis. Box plots represent the median and interquartile range and whiskers extend to the maximum or minimum values. Mann-Whitney U tests were used for comparisons (ns *p>*.05, \**p*≤ .05, \*\*\**p*≤.001, \*\*\*\**p* ≤.0001).

To establish a baseline of anti-RBD antibody binding by flow cytometry, convalescent samples from individuals recovered from acute SARS-CoV-2 infection were incubated with fluorescently labeled SARS-CoV-2 Wuhan-Hu-1 (Gene ID: 43740568) RBD-coated beads followed by fluorescently labeled secondary antibodies to specifically quantify anti-RBD IgG and IgA (Figures 1B-H). Due to the sensitivity of this assay, the quantifiable detection range is 10-50 ng/mL for RBD-specific antibodies (Sup. Figure 3) at dilutions of 1:10 for saliva (Figure 1B) and 1:900 for plasma (data not shown). For this reason, convalescent saliva (n=12) was serially diluted, establishing that a 1:10 ratio could quantifiably evaluate antibody levels up to a 500 ng/mL minimizing saturation without eliminating detectable signal in less concentrated samples (Figure 1B). Positive gates were drawn using pre-pandemic plasma (Sup. Fig. 4A) and separately on known negative saliva samples (Sup. Fig. 5A) and confirmed by convalescent plasma (Sup. Fig. 4B, Sup. Table 5) and saliva (Sup. Fig. 5B) samples. Anti-RBD IgA antibodies above pre-pandemic controls were detected in 89% of convalescent plasma samples analyzed (Figure 1C). Due to observed non-specific binding of salivary IgA to anti-RBD BioLegend beads in this study and by others,^42^ this data is not shown while further investigation into the specificity is undertaken. Anti-RBD IgG above the highest level detected in known negative samples was observed in 25% of convalescent saliva samples (Figure 1D) and in all convalescent plasma samples (n=19) analyzed (Figure 1C). As opposed to salivary IgA, salivary anti-RBD IgG binding is specific, and findings are consistent across all samples analyzed.

**Fig. 3:**
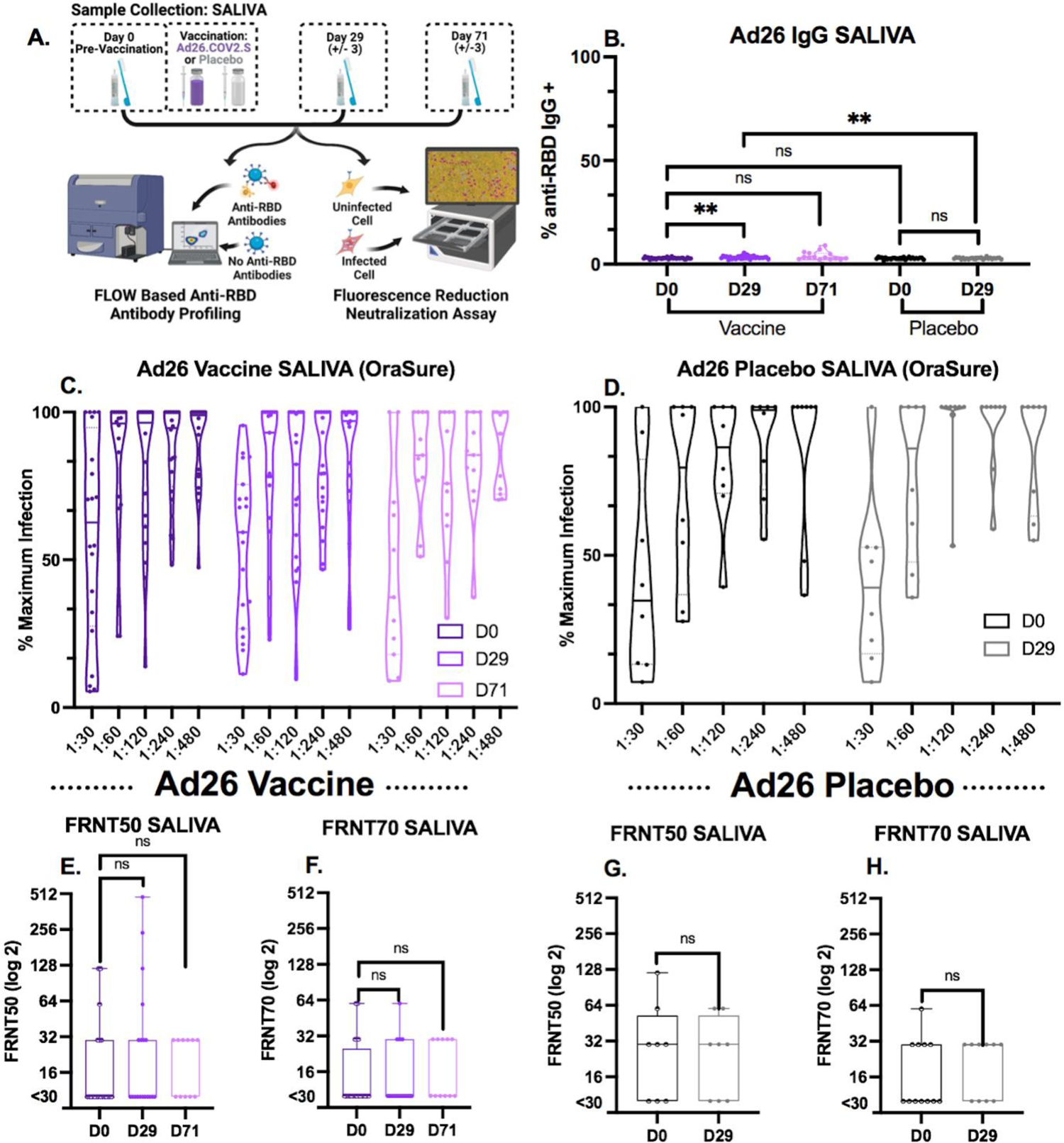
Ad26.COV2.S vaccinated individuals have very low levels of RBD-specific antibodies and lack differences in neutralization activity in saliva from vaccinated and placebo controls. **A:** Schematic of saliva collection timeline and downstream analysis. **B)** Levels of RBD-specific IgG in the saliva (OraSure collection device) of recipients of Ad26.COV2.S (n=31) or a placebo saline injection (n=27) measured via flow cytometry. Mann-Whitney U tests were used for comparisons (\*\**p* ≤.01). **C-D:** Neutralization activity in saliva (OraSure collection device) from D) individuals vaccinated with Ad26.COV2.S (n=20 for D0 and D29, n=11 for D71) or E) a placebo saline shot (n=8) represented as percent of maximum of triplicate rVSV-eGFP-SARS-CoV-2 control conditions. Each dot represents the fluorescent signal from rVSV-eGFP-SARS-CoV-2 infection from an individual patient in the assay. Any values over 100% of maximum infection were deemed as saturated and as such a threshold of 100% was set. Saliva samples were analyzed at two step dilutions ranging from 1:30-1:480. **E-H:** Fluorescence reduction in neutralization titer (FRNT) 50 and 70 of rVSV-eGFP-SARS-CoV-2 infection of HEK293-hACE2-mCherry expressing cells from saliva (OraSure collection device) of F-G) individuals vaccinated with Ad26.COV2.S (n=20 for D0 and D29, n=11 for D71) or H-I) a placebo saline shot (n=8). See Figure 1 for details on analysis. Box plots represent the median and interquartile range and whiskers extend to the maximum or minimum values. Mann-Whitney U tests were used for comparisons (ns *p>*.05).

**Fig. 4:**
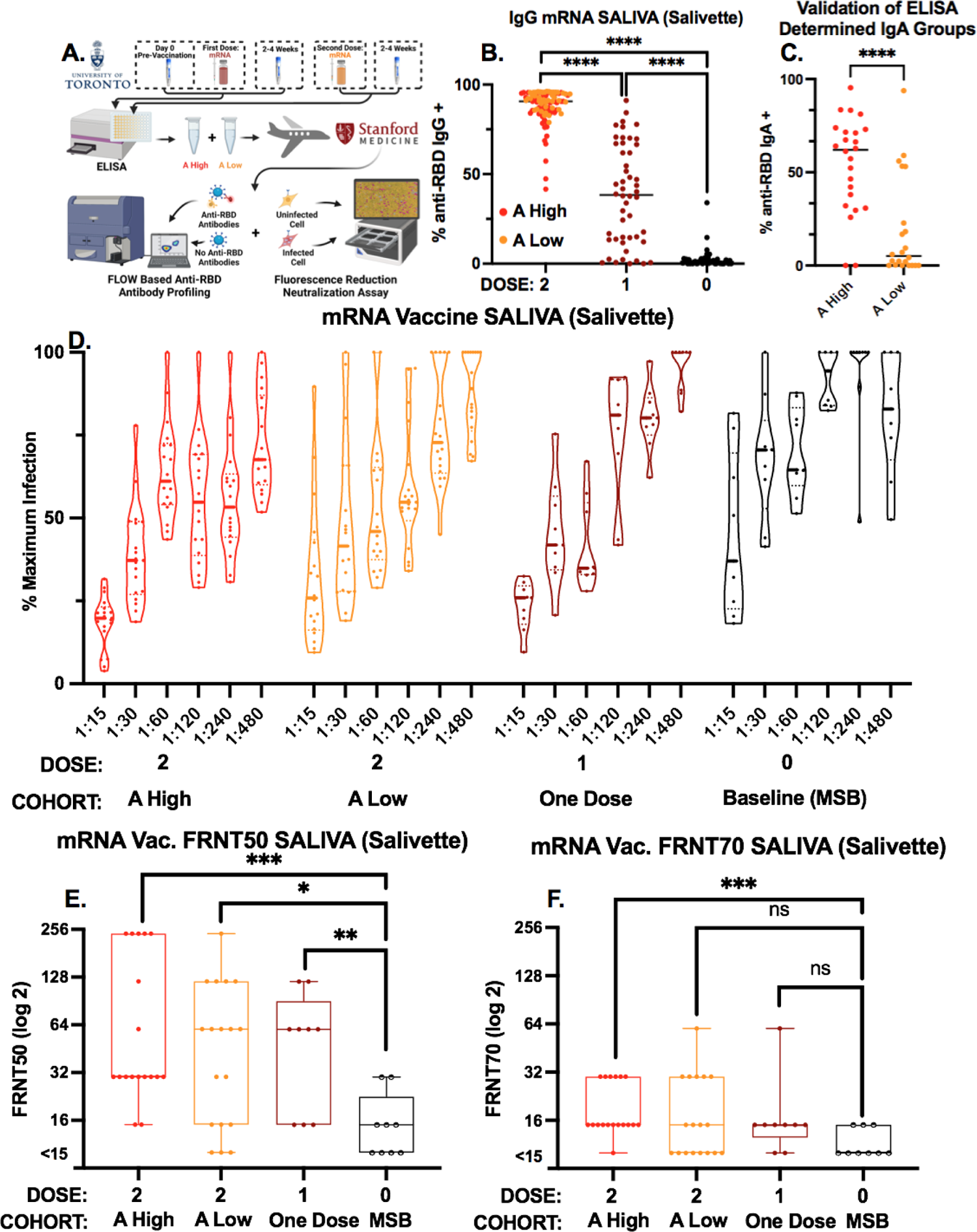
mRNA vaccines elicit a strong salivary RBD-specific antibody response and have significantly increased neutralization activity in the saliva. **A:** Schematic of sample exchange and the timeline of saliva collection and downstream analysis. **B:** Levels of RBD-specific IgG in the saliva of individuals having received either 2 (n=46), 1 (n=25), or 0 (n=25) doses of an mRNA vaccine measured via flow cytometric analysis. Saliva was collected using a Salivette device and evaluated by ELISA and characterized by their cutoff (2 standard deviations greater than the level of signal from pre-COVID-19 saliva samples). Mann-Whitney U tests were used for comparisons (\*\*\*\**p* ≤.0001). **C:** Levels of RBD specific IgA as determined by flow cytometry in the saliva of individuals who received 2 doses of an mRNA vaccine. A High and A Low correspond to Gommerman assigned designations that correspond to “A High” and “A Low” values derived from their ELISA. Mann-Whitney U tests were used for comparisons (\*\*\*\**p* ≤.0001). **D:** Neutralization activity of saliva (Salivette collection device) from individuals having received either 2 doses (n=24 “A High” and n=22 “A Low” per the University of Toronto assignment), 1 dose (n=25), or 0 (n=25) doses of an mRNA vaccine represented as percent of maximum of triplicate rVSV-eGFP-SARS-CoV-2 control conditions. Each dot represents the fluorescent signal from rVSV-eGFP-SARS-CoV-2 infection from an individual patient in the assay. Any values over 100% of maximum infection were deemed as saturated and as such a threshold of 100% was set. Saliva samples were analyzed at two step dilutions ranging from 1:15-1:480. **E-F:** Fluorescence reduction of neutralization titer (FRNT) E) FRNT50 and F) FRNT70 of rVSV-eGFP-SARS-CoV-2 infection of HEK293-hACE2 expressing cells from saliva (Salivette collection device) of individuals having received either 2 doses (n=24 “A High” and n=22 “A Low” per the University of Toronto assignment), 1 dose (n=25), or 0 (n=25) doses of an mRNA vaccine. See Figure 1 for details on analysis. Box plots represent the median and interquartile range and whiskers extend to the maximum or minimum values. Mann-Whitney U tests were used for comparisons (ns *p>*.05, \**p*≤ .05, \*\**p* ≤.01, \*\*\**p* ≤.001).

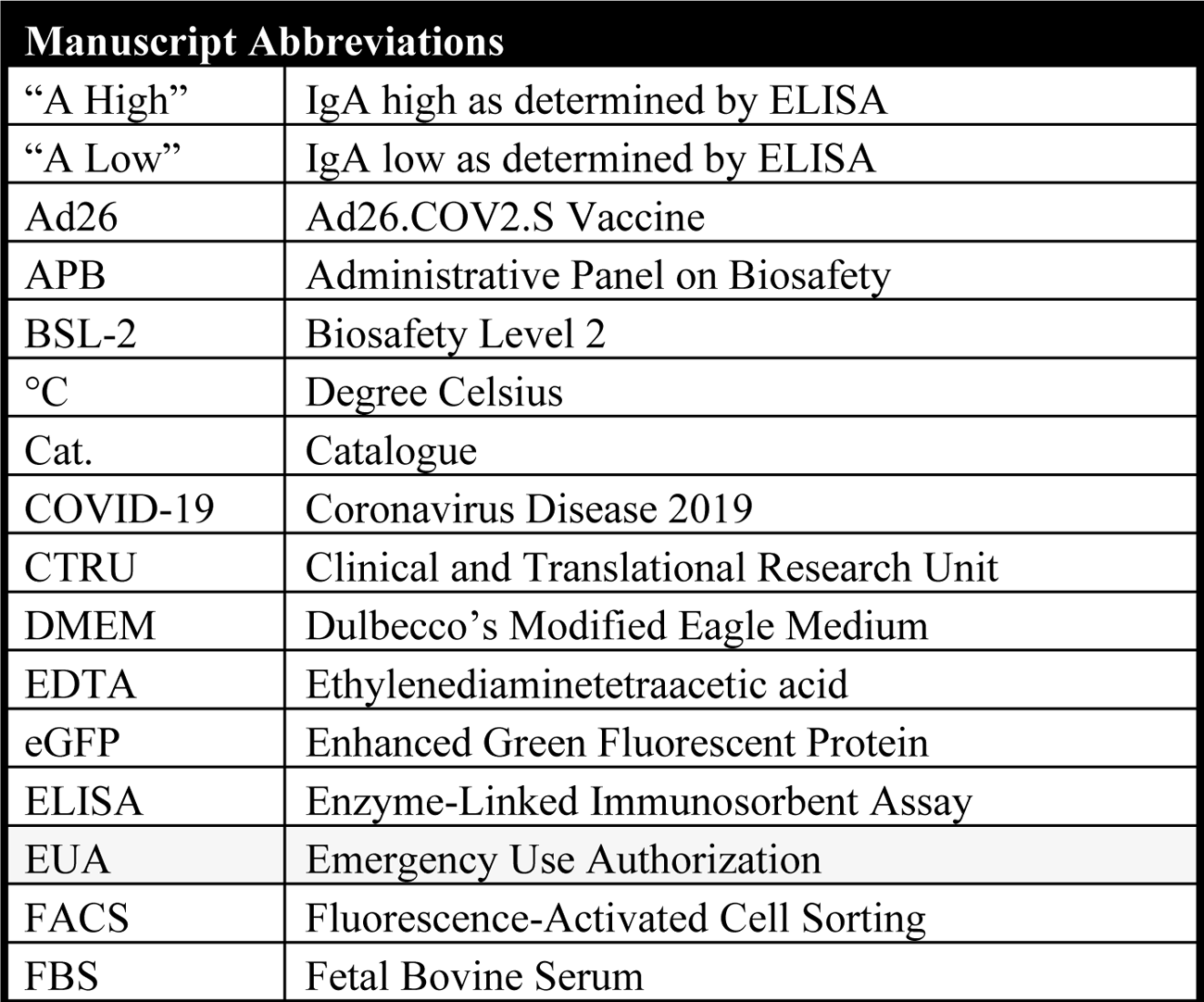
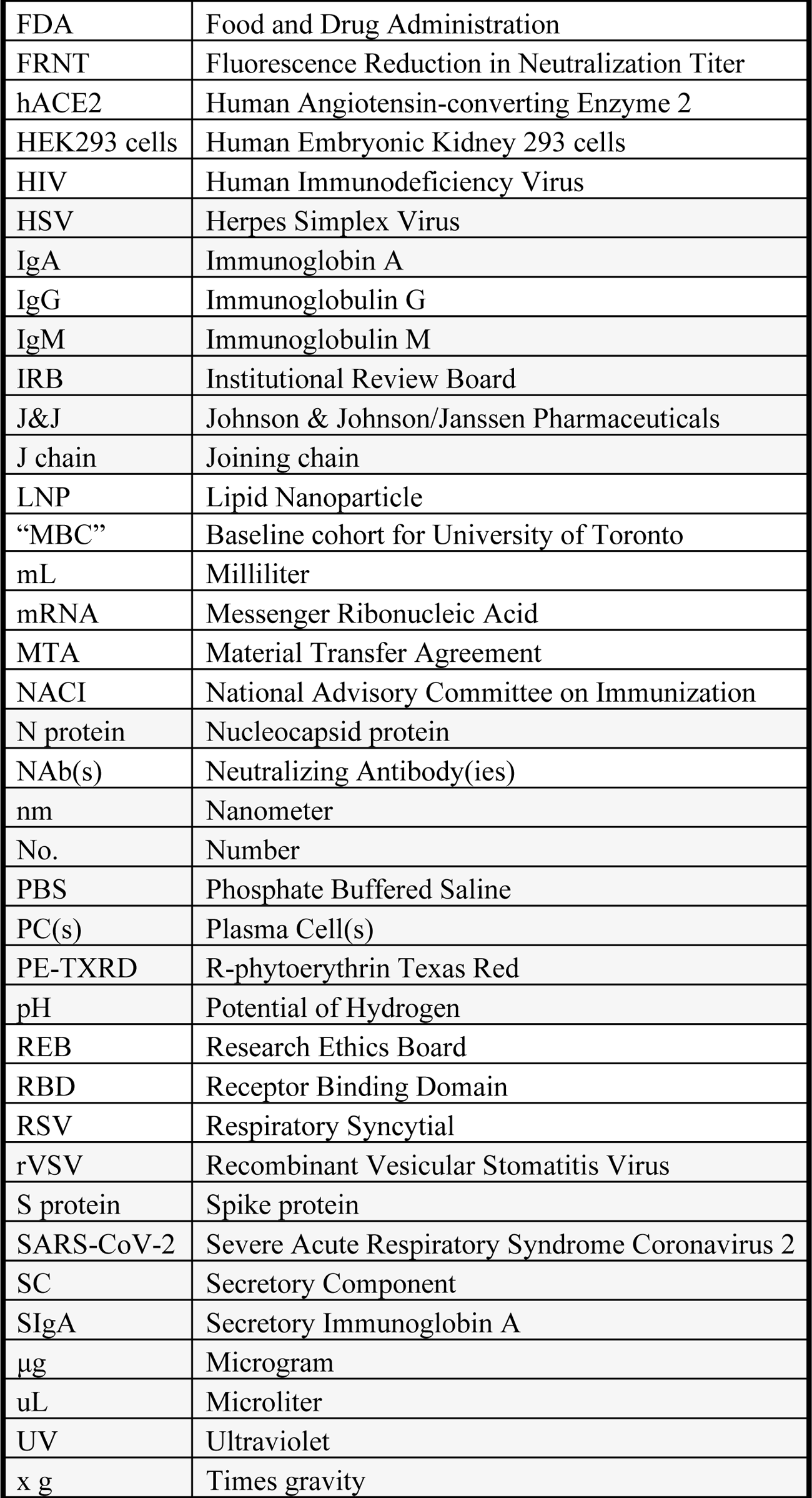

Neutralizing activity of SARS-CoV-2 antibodies in saliva and plasma were measured at two step dilutions (1:15-1:480 and 1:100-1:3200, respectively) following incubation with recombinant Vesicular Stomatitis Virus (rVSV)-eGFP-SARS-CoV-2-Spike (generously provided by Dr. Sean Whelan, Washington University, St. Louis (WUSTL)) in which the VSV-G protein was replaced with SARS-CoV-2-Spike protein (*43*). Virus supernatant was added to hACE2-mCherry expressing HEK293 cells (generously provided by Dr. Siyuan Ding, WUSTL) and infection was measured by green fluorescence over the course of 72 hours in an IncuCyte (Sup. Fig. 6).

Because the replication competent VSV that is expressing the spike protein from SARS-CoV-2 is labeled with eGFP, the level of infection is correlated with green fluorescence in the hACE2 expressing cells. Quantitative blocking can be assessed by the minimum dilution of a sample that has no more than 50% or 30% of the averaged green fluorescence signal of triplicate rVSV-eGFP-SARS-CoV-2 control wells. We included a serial dilution of a commercially available monoclonal antibody against RBD in every plate and graphed them as percentage of maximum of the averaged green fluorescence signal of triplicate rVSV-eGFP-SARS-CoV-2 control wells, as well as compared integrated fluorescence intensity values of each from all plates (Sup. Figure 7). We found that the percentage of maximum green fluorescence provided normalized values that could be fairly compared across different plates. We set two stringencies to quantify the green fluorescence reduction in neutralization titer (FRNT) of rVSV-eGFP-SARS-CoV-2 infection of HEK293-hACE2-mCherry cells, FRNT50 and FRNT70, which represent the minimum dilution that had 50% or 30%, respectively, of the average of the fluorescent signal of triplicate rVSV-eGFP-SARS-CoV-2 control wells as described in more detail in the methods.

Dilutions of 1:15 in saliva collected in OraSure devices have been omitted from analyses as the preservative solution alone resulted in rapid cell death (Sup. Figure 8). With much higher levels of antibody in the plasma as compared to saliva, at a dilution of 1:800 green fluorescence of the rVSV-eGFP-SARS-CoV-2 infection of HEK293-hACE2-mCherry cells was still below 50% of maximum compared to uninfected controls (Figure 1E). In saliva samples collected by OraSure, a dilution of 1:30 reduced infection as compared to known negative samples (Figure 1F). FRNT50 was significantly different in convalescent saliva when compared to controls, while FRNT50 and FRNT70 showed highly significant neutralizing activity in the plasma of convalescent individuals when compared to controls (Figures 1G, 1H).

We carried out an observational study to investigate the mucosal and systemic antibody responses induced in response to vaccination. One cohort of 64 participants (Sup. Table 1; D29 placebo=33; vaccine=31) were enrolled in the J&J Phase 3 ENSEMBLE trial of the Ad26.COV2.S vaccine and were studied via longitudinal sampling of plasma (Figure 2A) and saliva (Figure 3A) post-vaccination. This was not a randomized prospective study. There are many issues in designing a prospective study, which would have taken into account a number of characteristics of the population being followed which were not accounted for in this study.

Cohorts and subgroups were not randomized or case controlled. Study staff was blinded to the vaccination status of participants. Sample unblinding occurred only after samples were analyzed for anti-RBD IgG and IgA by flow cytometry. From this cohort, two participant sample sets from the placebo group were omitted from our analysis due to natural infection or because they opted for mRNA-1273 or BNT162b2 vaccinations, which became available to the public during our sampling period (Sup. Figures 9, 10 and 11). In parallel to the recruitment and collection of this cohort, a separate cohort receiving EUA mRNA vaccines (n=23) were longitudinally sampled over the course of receiving 2 doses of vaccine were collected using the same collection device (Sup. Table 5, Sup. Figure 12). Finally, a cohort of single and 2 dose mRNA vaccine recipients was collected using the Salivette saliva collection device by the Gommerman Lab at the University of Toronto and provided to Stanford through Material Transfer Agreement for antibody evaluation and pseudotyped virus neutralizing potential in our fluorescence reduction of neutralizing titers assay.

As baselines for antibody levels were established using COVID-19 convalescent saliva and plasma as compared to pre-pandemic plasma and known negative saliva, we sought to evaluate the antibody responses produced in response to vaccination beginning with the previously described Ad26.COV2.S cohort. By day 29 (+/- 3 days) post Ad26.COV2.S vaccination, levels of anti-RBD specific IgG and IgA in the plasma were detectable by flow cytometric analysis (Figures 2B, 2C). Of the vaccinated group (n=31), 67.7% generated anti-RBD IgG antibodies and 29% generated anti-RBD IgA levels that were detectable above the highest levels in the placebo group (n=27). The placebo group exhibited no significant changes in antibody titers from day 0 to day 29 (Figure 2B, 2C). In a subset of the Ad26.COV2.S vaccine cohort, a third sample set was collected at day 71 (+/- 3 days) post vaccination (n=14) where anti-RBD IgG plasma titers remained detectable in 33% of vaccine recipients, while detectable anti-RBD IgA lost statistical significance. Flow cytometric values saturate at 0.05 μg/mL (Sup. Figure 3), so all plasma were diluted 1:900 in phosphate buffered saline (PBS) to enable quantification of up to 45 μg/mL before saturation. Increased anti-RBD IgG titers observed by day 29 trend with increased neutralizing activity at both day 29 and day 71 post vaccination (Figure 2D) as quantified by FRNT50 and FRNT70 (Figures 2F, 2G) when compared to no change in placebo group (Figures 2E, 2H, 2I).

Relative to the plasma of Ad26.COV2.S vaccine recipients, we found small but significant increases in levels of anti-RBD IgG antibodies at day 29 in the saliva of Ad26.COV2.S vaccine recipients compared to placebo recipients (Figure 3B). This observation is also seen when comparing anti-RBD IgG from day 0 and day 29 for vaccinated individuals using their pre-vaccination time-point (Figure 3B). At the day 71 timepoint, the samples were not statistically significant from day 0, but this could be due to a lower n of 14 individuals at the later timepoint and a wider variability of responses, including several below the limit of detection. A separate cohort of mRNA vaccinated individuals (n=23) who provided longitudinal saliva samples using the same OraSure collection device, shows increasing anti-RBD IgG from day 0 to at least day 60 (Sup. Figure 12), indicating that the findings are due to differences in vaccination design.

Analysis of anti-RBD IgA showed a lack of specificity, but with equal amounts of background signal across all samples from before or after vaccination in placebo and vaccine groups (data not shown). Furthermore, the saliva samples from the Ad26.COV2.S vaccinated group exhibited no difference in neutralizing activity (Figure 3C) by either FRNT50 or FRNT70 quantification (Figures 3E, 3F) from day 0 to day 29 or day 71 post vaccination, or in comparison to the placebo group (Figures 3D, 3G, 3H). Because of the activity of factors in the saliva or the OraSure preservative that impact the rVSV-eGFP-SARS-CoV-2 assay at the 1:15 (cell toxic) and 1:30 (high noise) dilutions there is a significant amount of noise at the lowest dilutions which limits the sensitivity of the assay (Sup. Figure 8).

In collaboration with the Gommerman lab at the University of Toronto, saliva samples that were collected using Salivettes from BNT162b2 or mRNA-1273 vaccine recipients (Sup. Table 2) were shared with Stanford and independently evaluated by previously described flow cytometric analysis (n=94) for anti-RBD IgG and IgA antibody titers and by live-virus fluorescence reduction neutralizing activity (n=54) (Figure 4A). Samples were collected 2-4 weeks post vaccination and evaluated for anti-RBD and anti-Spike antibodies by Enzyme Linked Immunosorbent Assay (ELISA). From Gommerman ELISA analysis, the samples were divided into two groups where the number of vaccine doses administered was either 1 dose (n=50) or 2 doses (n=44). The single dose vaccine group included a “baseline” sample set to act as control and the 2-dose vaccine sample set was further divided into two groups where anti-RBD IgA titers measured by the Gommerman Lab using an ELISA were designated as either “IgA High” or “IgA Low” referred to from this point and in the figures as “A High” and “A Low”.

When the Gommerman Lab samples were analyzed using flow cytometry, we found that statistically significant titers of anti-RBD IgG averaging over 500 ng/mL were detectable in both cohorts of single and double vaccine doses, with Gommerman designated “A High’’ and “A Low” (*44*), of individuals who had received two doses of vaccine clustering together at saturated levels of anti-RBD IgG (Figure 4B). Single dose RBD-specific IgG antibody titers were similar to convalescent saliva (Figures 1D, 4B). Evaluation of anti-RBD IgA titers during blinded flow cytometry profiling of these samples showed comparable findings of IgA levels overall as the “A High” and “A Low” ELISA designations. With only slight variations between the antibody detection methods, the signal detected using BioLegend RBD beads in several of the ELISA designated “A Low” is most likely due to non-specific salivary IgA binding to the surface of the RBD beads (Figure 4C), which is consistent with high salivary IgA binding on beads in control conditions seen in similar multiplexed assays (*45*). In the ELISA assays used by the Gommerman lab to quantify salivary anti-RBD levels, samples were pre-adsorbed with streptavidin to eliminate non-specific streptavidin binding IgA in saliva as previously described (*33*). The lack of non-specific binding in pre-adsorbed samples is reflected in their low level of IgA signal derived from pre-COVID-19 saliva (data not shown, please see accompanying manuscript) (*44*). Maximum infection and FRNT50 quantification of neutralizing activity in 1-dose and “A Low’’ 2-dose groups all had significantly higher neutralizing activity compared to Baseline (Figures 4D, 4E), as well as comparable neutralizing activity to convalescent saliva collected by OraSure (Figures 1G, 1H). Of all the saliva samples from vaccinated individuals that we examined, the only cohort to reach statistical significance at the stringent quantification of FRNT70 was the two-dose mRNA-1273/BNT162b2 vaccine group provided by the University of Toronto, who also had high levels of IgA in the saliva (designated “A High”) by ELISA (Figure 4F).

## DISCUSSION

Currently, our best measure of defense against SARS-CoV-2 is vaccination, which can be further supplemented with other preventative interventions. All three EUA vaccines have evidence of “breakthrough infections” or infections in fully vaccinated individuals (*46, 47*). The amount of mucosal immunity needed to prevent infection and whether any vaccines have achieved these protective levels remains unknown. If intramuscular vaccinations do not provide universal or complete resistance of oronasopharyngeal infection by SARS-CoV-2, breakthrough infections could occur without the development of serious systemic disease; such individuals who are vaccinated could still harbor and spread infection to others who are susceptible.

For maximum protection against infection and variants with increased transmissibility, inducing mucosal antibodies is important. Existing vaccines aimed at inducing local mucosal antibodies include intranasal spray (FluMist) for influenza (*48*) and oral drops for rotavirus (RotaTeq/Rotarix) (*49*), typhoid (Vivotif) (*50*) and polio (oral poliovirus vaccine, OPV) (*51*). A monoclonal antibody (MAb362) sIgA showed the ability to neutralize live SARS-CoV-2, while MAb362 IgG, even at the highest tested concentration, did not neutralize (*52*). Notably, nasal delivery of IgM has been shown to offer broad protection from SARS-CoV-2 variants (*53*). Additionally, a single dose intranasal ChAd-SARS-CoV-2 S vaccine in mice induces durable and neutralizing IgG and IgA antibodies, which are effective in protecting against new variants of concern (*54*).

Importantly, our study was observational and not structured to directly compare between SARS-CoV-2 vaccines, with significant differences in participant cohorts, timepoints, and saliva collection devices. Based on our data, natural infection induces production of anti-RBD IgG and IgA with neutralizing activity in the saliva and plasma of convalescent individuals. All vaccines evaluated in our study produce robust systemic immune responses in most individuals, with higher levels of anti-RBD antibodies and neutralizing activity following two doses of mRNA vaccination as compared to a single dose of Ad26.COV2.S. As reported previously, the mRNA vaccines, BNT162b2 or mRNA-1273, induced high levels of RBD-specific IgG antibodies in the saliva (*36–39, 45*), at concentrations over 500 ng/mL after the second dose. We found that these vaccines generated neutralizing activity in the saliva comparable to natural infection, with peak levels after the second dose in those individuals which had high levels of IgA in their saliva as determined by ELISA. The SPIKE and RBD binding IgA antibodies in these samples were shown to have secretory component as would be expected of dimeric IgA in the saliva (*44*), and importantly, dimeric IgA has been shown to be a more potent neutralizer than either IgG or monomeric IgA (*55*).

The detection of high amounts of IgG in the saliva opens the question of how that IgG arrived there. Detectable levels of circulating spike protein after mRNA vaccine have been shown to persist for several days post vaccination (*56*), and one possibility which some of the authors find worthy of speculation is if this could induce a mucosal immune response, which has not historically been reported with other formulations of intramuscular vaccination. Alternatively, salivary IgG may be a result of antibody titers, as a generally held assumption is that passive transudation of neutralizing antibodies accounts for the high levels of mucosal IgG. Testing this assumption directly is important, as the high plasma titers could give rise to IgG by bleeding contamination. Future work could examine the extent to which monoclonal salivary IgG is detectable following high dose intravascular injection with a monoclonal antibody that could be detected with reagents such as an anti-idiotype antibody based test. However generated, such a mechanism could have important implications for protection against other viruses that infect mucosal surfaces, while also proving difficult to develop effective vaccinations for, including Respiratory Syncytial Virus (RSV), Herpes Simplex Virus (HSV), and Human Immunodeficiency Virus (HIV) (*57*). If intramuscular mRNA vaccines can consistently catalyze systemic and mucosal immune responses to SARS-CoV-2, we suggest incorporating these approaches to immunization.

The innate and adaptive immune responses are extremely complex and we do not know all of the elements that will lead to a protective immune response. In addition to neutralizing antibodies, cellular immunity provides significant and durable protection from infection. Ad26.COV2.S has been shown to induce strong T cell responses (*6, 58*), which could potentially overcome the absence of local antibodies. In this study, we did not measure cell mediated immunity to virus infected cells. Nevertheless, vaccinated recipients with lower antibody titers and neutralizing activity in the saliva suggest that there could be potential value in boosting immunity with a subsequent dose of an appropriate vaccine.

### Limitations

Researchers are still learning about the biology around these different vaccine formulations, which may elicit distinct types of antibody responses and specifically salivary antibodies. Furthermore, researchers are still learning about the immune components in saliva. For example, the method of assay may require special handling steps such as the pre-adsorption step with materials not containing the RBD or spike protein targets to reveal specific antibodies to the viral targets, as shown by the Gommerman lab ELISA assay(*33, 44*) and others (*45*). One limitation of our observational study is that the extent that salivary antibodies contribute to vaccine efficacy remains unknown, and further research should consider how salivary antibodies may explain some differences in observed vaccine efficacy. Antibodies are only one facet of a complex immune response, and therefore an important limitation of our study is that we did not investigate T-cell responses, which are seen to provide immune protection following vaccination even in the absence of protective antibodies. We were surprised to find such high levels of IgG isotype antibodies in the saliva of mRNA vaccinated individuals. In our limited testing, we found salivary IgA anti-RBD in response to natural infection, but not in all mRNA vaccinated individuals. Humans practice oral hygiene and perturbations to the mucosa that experimental animals do not. The elements of oral hygiene can lead to injury where individuals with high titer serum antibodies may have contamination of salivary fluids with systemic antibodies. Given the high levels of circulating antibodies generated from vaccination, as evident from the 1:900 dilution of plasma samples, reported saliva results could be skewed. Further, saliva samples obtained using the Salivette collection device show resolution at 1:15 dilution in the FRNT assay, whereas OraSure collected samples resolve at the next dilution step of 1:30, which impacts the lower limit of detection. Finally, while the n for this study is small, cross validation with the Gommerman lab at the University of Toronto warrants strong consideration for the results reported herein.

## METHODS

### ETHICS STATEMENT

The Stanford University, School of Medicine Institutional Review Board (IRB) granted approval for recruiting SARS-CoV-2 vaccine trial participants and EUA vaccine recipients to this study for saliva and blood collection and for studying the antibody response to SARS-CoV-2 antigens in those samples (study number 57277 and 55689). The University of Toronto Research Ethics Board (REB) provided approval for Stanford to conduct antibody analysis of saliva samples to SARS-CoV-2 antigens for samples collected under study number 23901.

### EXPERIMENTAL MODEL AND SUBJECT DETAIL

#### Study Participants

Participants in this study were either sole participants of this IRB 57277 study or co-enrolled in the Johnson & Johnson/Janssen Pharmaceuticals (J&J) Phase 3 ENSEMBLE trial (Sup. Table 1, 3 and 5). Additional participants were recruited through social media posts, news media articles citing this research and/or word of mouth. Those individuals recruited directly to this study were requested to self-report if they had or intended to receive a vaccination against SARS-CoV-2 including BNT162b2, mRNA-1273, or Ad26.COV2.S, and/or if they had recovered from infection with SARS-CoV-2. All participants in this study were between the ages of 18-85, reported their biological sex assigned at birth as either male or female, were not pregnant at the time of sample collection, and self-attested to being healthy at the time of sample collection.

### Human Samples

#### PLASMA

COVID-19 convalescent plasma samples were purchased from the Stanford Blood Center. Samples were obtained from individuals who exhibited mild symptoms of acute COVID-19, had a positive RT-PCR result for SARS-CoV-2, and who had complete resolution of symptoms followed by a negative RT-PCR test at least 14 days after initial symptom onset (Sup. Table 3) Pre-pandemic plasma samples were obtained in 2018 as part of another study, IRB 46112, prior to the circulation of SARS-CoV-2 in the global population. Participants in this study, IRB 46112, consented for samples to be saved for future research purposes (Sup. Table 6).

Plasma and saliva obtained from participants co-enrolled in this study and the J&J Phase 3 ENSEMBLE trial (Sup. Table 1) were collected in parallel at the time of visit at the Stanford Clinical and Translational Research Unit (CTRU). These participants consented to provide samples at their initial visit prior to receiving vaccination or placebo and during their first follow-up visit approximately 29 days (+/- 3 days) post-vaccination. In a subset of participants, a third sample set was collected at their second follow-up visit approximately 71 days (+/- 3 days) post-vaccination.

#### SALIVA

Saliva was collected from study participants using the OraSure Oral Specimen Collection Device (OraSure®, Cat. No. 3001-2870) which collects saliva produced by the salivary glands (Sup. Tables 1, 4 and 7). The absorbent paddle of the OraSure Collection Device is gently brushed against the gums 1-2 times and left in place between the gum and cheek for 2-5 minutes. The absorbent paddle is then removed from the oral cavity and placed in a provided collection tube containing preservative. The collected sample was then stored at 4 °C for up to 21 days prior to biobanking. In some cases, the completed collection devices remained at room temperature during shipping and were transferred to 4 °C upon arrival at Stanford. A subset of samples were collected by the Stanford Nadeau lab (IRB 55689) study, which longitudinally samples various tissues including saliva and plasma from patients recovered from acute SARS-CoV-2 infection. A subset of saliva samples from individuals who received zero doses, a single dose or two doses of either the BNT162b2 or mRNA-1273 vaccine that were analyzed in this study were collected by the Gommerman lab at the University of Toronto (REB 23901) and were shared with Stanford under a Material Transfer Agreement (MTA) agreement (Sup. Table 2). Saliva samples from this cohort were collected using Salivette® tubes (Sarstedt, Numbrecht, Germany), a collection system which consists of a cotton ball which participants chew for exactly three minutes and place into a tube, which is then placed into a larger outer tube. The entire system is spun in a centrifuge at 1000 times gravity (x g) for five minutes at room temperature. The inner tube contains a hole at the bottom, which allows all the saliva absorbed by the cotton ball to filter into the larger outer tube. The total saliva volume from each participant was then aliquoted and stored at –80°C. Given that these samples were collected from vaccinated participants who reported no symptoms of COVID-19 infection, we did not conduct any measures for viral inactivation.

### Biobanking Procedure

Prior to sample processing, all samples were ultraviolet (UV) irradiated at 254 nanometers (nm) for 15 minutes. Heat inactivation of samples resulted in loss of signal. All samples were handled and processed according to Stanford’s Biosafety Level 2+ (BSL2+) guidelines for the handling of infectious human tissues under APB 2970.

Whole blood samples were collected in lavender top ethylenediaminetetraacetic acid (EDTA) coated tubes (Fisher Scientific, Cat. No. 02-683-99C) and centrifuged at 1500 x g for 15 minutes on the same day as collection. Serum was aliquoted into 1.5 mL tubes as assay appropriate volumes and stored at –80 °C. One aliquot from the red blood cell pellet was also taken at this time and stored at –80 °C.

Collected OraSure sample devices with cap were spun at 1500 x g for 15 minutes in secondary containment in open-top thin wall ultra-clear tubes (Beckman Coulter, 344058) to remove preservative containing sample. Spun samples were aliquoted and stored at –80 °C for long-term storage.

## METHOD DETAILS

### Flow Cytometric Analysis

For flow cytometric analysis, saliva samples were diluted in 1x phosphate buffered saline (PBS) at 1:10, and plasma samples were diluted in 1x PBS at 1:900. These samples were incubated overnight at 4 °C on a shaker in 96-well V-bottom plates with BioLegend 13x RBD beads (BioLegend, Cat. No. 741136) at the manufacturer’s recommended concentration per reaction. Samples were washed and then stained with bulk IgG in Alexa Fluor 488 (Southern Biotech, Cat. No. 9042-30) and bulk IgA in R-phycoerythrin Texas Red (PE-TXRD) (Southern Biotech, Cat. No. 2050-07) for 20 minutes on ice. Samples were washed twice and then resuspended in flow cytometry buffer (2 mM EDTA, 2% Fetal Bovine Serum (FBS), 1x PBS pH 7.4). Samples were analyzed for antibody reactivity against RBD by flow cytometry using a BD LSRFortessa Cell Analyzer. Gating scheme is shown in supplemental figures 4 and 5 comparing percentage antibody bound to RBD beads and known concentrations of anti-RBD IgG quantified.

### Fluorescent Reduction of Neutralization Titers

#### VIRUS

The recombinant Vesicular Stomatitis Virus (rVSV) used in this assay was a gift from Dr. Sean Whelan, (WUSTL). The rVSV expresses enhanced Green Fluorescent Protein (eGFP) in place of the glycoprotein and has been further engineered to express the full-length Wuhan-Hu-1 Spike protein. rVSV-eGFP-SARS-CoV-2-S was propagated on MA104 cells (courtesy of Dr. Siyuan Ding, WUSTL) as previously described (*43*). MA104 cells were maintained in Medium 199 (Gibco, Cat. No. 11150067) supplemented with 10% FBS and 1% Penicillin/Streptomycin (Fisher Scientific, Cat. No. 15-140-163). After visible cytopathic affect, supernatant was filtered, aliquoted and stored at –80 °C.

#### CELLS

Human Embryonic Kidney (HEK) 293 cells were engineered to encode human angiotensin converting enzyme 2 (hACE2 in the pDEST-mCherry vector) (courtesy of Dr. Siyuan Ding, WUSTL) as previously described (*59*). HEK293-hACE2-mCherry cells were cultured in Gibco Dulbecco’s Modified Eagle Medium (DMEM) formulation containing glucose, L-glutamine and sodium pyruvate (Gibco, Cat. No. 11995065) with 1x Penicillin/Streptomycin. Geneticin Selective Antibiotic (G418) (Gibco, Cat. No. 10131035) was added at a concentration of 500 μg/mL to maintain hACE2-mCherry expression. Cells were grown in 5% carbon dioxide (CO2) at 37 °C and passaged every 3 days using Versene solution (Gibco, Cat. No. 15040066).

#### ASSAY

HEK293-hACE2-mCherry cells were seeded at a density of 25,000 cells per well in a 96-well, flat-bottom tissue culture coated plate. Outer rows were avoided to reduce assay variations resulting from edge effect in the IncuCyte. In a separate 96 well plate, samples were serially diluted and incubated with 50 μL of rVSV-eGFP-SARS-CoV-2-S for 2 hours at 37 °C in 5% CO2. Each sample plate included a dilution of anti-RBD antibody (Invitrogen, Cat. No. 703958) of 10 μg/mL, 5 μg/mL, 1 μg/mL, 0.5 μg/mL, 0.1 μg/mL, and 0.05 μg/mL. After incubation, the mixture of sample and rVSV-eGFP-SARS-CoV-2-S was transferred to the plated HEK293-hACE2-mCherry cells at a 1:1 ratio of culture media to virus/sample suspension. Plates loaded in the IncuCyte were imaged every 3 or 4 hours for a total of 72 hours with 4 scans per sample well to visualize neutralization. Representative plate layouts for each cohort presented (Figures 1E & F, 2D & E, 3C & D, and 4D) of IncuCyte generated integrated fluorescence intensity levels at all time points across representative plates with samples presented in this paper show the per-plate controls as well as the samples green fluorescence (Sup. Figures 13-15).

## QUANTIFICATION AND STATISTICAL ANALYSIS FRNT50 and FRNT70

To quantitatively determine assay sensitivity, normalized anti-RBD curves from every neutralization assay performed in this study were plotted (Sup. Figure 10A). This assay is sensitive down to 5 μg/mL of neutralizing antibodies. Due to natural variance in the total integrated intensity of anti-RBD control curves included in each plate (Sup. Figure 10B), each plate was normalized either to the mean of the rVSV-eGFP-SARS-CoV-2-S supernatant controls or to the 0.05 μg/mL anti-RBD antibody. Normalization to 0.05 μg/mL anti-RBD was performed only if division by the triplicate rVSV-eGFP-SARS-CoV-2 control conditions resulted in loss of a sigmoidal shape of the anti-RBD curve. Any values over 100% of maximum infection were deemed as saturated and as such a threshold of 100% was set. FRNT50 and FRNT70 represent the minimum dilution that had 50% or 30%, respectively, of the average of the fluorescent signal of triplicate rVSV-eGFP-SARS-CoV-2 control wells.

### KEY RESOURCES TABLE

| REAGENT or RESOURCE | SOURCE | IDENTIFIER |
| --- | --- | --- |
| **Antibodies** |  |  |
| Anti-RBD antibody | Invitrogen | Cat. No. 703958 |
| Bulk IgM in Pacific Blue | BioLegend | Cat. No. 314514 |
| Bulk IgG in Alexa Fluor 488 (AF488) | Southern Biotech | Cat. No. 9042-30 |
| Bulk IgA in R-phycoerythrin Texas Red (PE-TXRD) | Southern Biotech | Cat. No. 2050-07 |
| **Bacterial and virus strains** |  |  |
| Recombinant Vesicular Stomatitis Virus (rVSV) | Dr. Sean Whelan, University of Washington (St. Louis) |  |
| **Biological samples** |  |  |
| **Chemicals, peptides, and recombinant proteins** |  |  |
| **Critical commercial assays** |  |  |
| BioLegend 13x RBD beads | BioLegend | Cat. No. 741136 |
| BioLegend N beads | BioLegend | Cat. No. 741137 |
| **Deposited data** |  |  |
| **Experimental models: Cell lines** |  |  |
| MA104 cells and HEK293 cells engineered to encode hACE2 in the pDEST-mCherry vector | Dr. Siyuan Ding, Washington University, St. Louis |  |
| Engineered rVSV-eGFP-SARS-CoV-2-Spike | Dr. Sean Whelan at Washington University, St. Louis |  |
| Experimental models: Organisms/strains |  |  |
| Oligonucleotides |  |  |
| Recombinant DNA |  |  |
| Software and algorithms |  |  |
| BD FACSDiva | BD Biosciences |  |
| Other |  |  |
| ` | OraSure® | Cat. No. 3001-2870 |
| Open-top thin wall ultra-clear tubes | Beckman Coulter | Cat. No. 344058 |
| Purple top EDTA tubes | Fisher Scientific | Cat. No. 02-683-99C |
| Medium 199 | Gibco | Cat. No. 11150067 |
| 1% Penicillin/Streptomycin | Fisher Scientific | Cat. No. 15-140-163 |
| Geneticin Selective Antibiotic (G418) | Gibco | Cat. No. 10131035 |
| Gibco Dulbecco's Modified Eagle Medium (DMEM) formulation containing glucose, L-glutamine and sodium pyruvate | Gibco | Gibco, Cat. No. 11995065 |
| Versene | Gibco | Cat. No. 15040066 |
| BD LSRFortessa | BD Biosciences | 649225 |

### RESOURCE AVAILABILITY

#### Lead Contact

Correspondence and requests for materials should be addressed to and M.C.T and I.L.W.

### Materials Availability Data and Code Availability

The code generated during this study is available at: https://github.com/georgieNahass/polarBarPlotsCovid/

## Supporting information

research reporting checklist

## ACKNOWLEDGEMENTS

Additional author members of the Stanford COVID-19 Biobank Study Group include: Jennifer A. Newberry, James V. Quinn, Rosen Mann, Anita Visweswaran, Elizabeth J. Zudock, Jonasel Roque, Hena Naz Din, Komal Kumar, Kathryn Jee, Brigit Noon, Jill Anderson, Bethany Fay, Donald Schreiber, Nancy Zhao, Rosemary Vergara, Julia McKechnie, Aaron Wilk, Lauren de la Parte, Kathleen Whittle Dantzler, Maureen Ty, Nimish Kathale, Arjun Rustagi, Giovanny Martinez-Colon, Geoff Ivison, Ruoxi Pi, Maddie Lee, Rachel Brewer, Taylor Hollis, Andrea Baird, Michele Ugur, Drina Bogusch, Georgie Nahass, Kazim Haider, Kim Quyen Thi Tran, Laura Simpson, Andrea Fernandes, Neera Ahuja, James Krempski. The authors wish to thank members of the Stanford University, School of Medicine, Weissman, Blish, and Nadeau labs, for helpful advice, discussions, and reagents. We would like to thank the members of Gommerman lab of the University of Toronto for collaborating on this study. M.C.T wishes to thank Tomer Tal, R.S.S wishes to thank Reese Shulman, E.C.S. wishes to thank Gabe Sanders, for their incredible support and sharing of childcare responsibilities to enable them to conduct this research and work on this manuscript. We would like to also thank Tomer Tal for input on statistical analysis. We would like to thank the lab of Dr. Sean Whelan at WUSTL, for their gift of rVSV-eGFP-SARS-CoV-2, Dr. Siyuan Ding at WUSTL, for mCherry labeled hACE-2 expressing HEK293 cells, Kevin Ng for discussion of neutralization assays, Dr. Thiago Carvalho for assistance with the discussion, Dr. Katherine Dantzler for antibodies in a desperate pinch, and Dr. Kim Hasenkrug and Dr. Lara Meyers at the National Institutes of Health (NIH) Rocky Mountain Labs for longterm discussions of the general topic of immune responses to infectious agents, especially viruses, and the role of macrophages in immunity to viruses, both local and systemic The authors are also grateful to individuals at the University of Toronto who collected and processed samples for these studies. These include Keelia Quin de Launay, Alyson Takaoka, Julia Garnham-Takaoka and Christina Fahim. Dr. Timothée Bruel, Dr. Caroline Goujon, and Dr. Jacob Yount sent us reagents that were not used in this manuscript but greatly assisted in the development of the assays that were used in this manuscript and we are very grateful. Schematic Figures 1A, 2A, 3A, 4A, and Supplemental Figure 1 were generated in BioRender. Research reported in this publication was supported by: The Fairbairn Family Foundation, The Stanford SPARK Program, Virginia and D.K. Ludwig Fund for Cancer Research. M.C.T., S.G. and P.H. were supported by the Bay Area Lyme Foundation, M.C.T was also supported by Robert J. Kleberg, Jr. and Helen C. Kleberg Foundation. G.R.N. and G.B. were supported by the Younger Family Foundation. R.S.S. was supported by Fairbairn Family Foundation. The funders had no role in study design, data collection and analysis, decision to publish, or preparation of the manuscript.

## AUTHOR CONTRIBUTIONS

G.R.N., R.S.S., M.C.T, and G.B. conducted experiments, prepared figures and also wrote portions of the manuscript. G.R.N. also performed neutralization assay data analysis and R.S.S. performed flow cytometry assay data analysis. G.R.N., G.B., C.A.B. and K.H. coordinated sample collection and storage and managed a sample database. R.S.S. created biobank sample database. R.B., G.R.N, R.S.S, M.C.T, G.B., and K.T. recruited and collected samples from individuals receiving vaccination. E.D., I.C., T.S., A.S.L., M.M. coordinating clinical sample collection and storage. R.S.S. wrote and managed IRB 57277. G.R.N. and G.B. created figures. Y.Y.Y. helped with editing the manuscript, coordinating sample collection and generating protocols. S.D.G. performed phlebotomy and sample intake, G.M-A and R.B. helped with sample intake. P.S.H helped with sample intake and IncuCyte use. K.C.N. provided samples and funding. E.C.S. helped manage sample database and wrote portions of the manuscript. I.L.W. and M.C.T. oversaw the research, helped design and analyze experiments as well as wrote portions of the manuscript.

## Supplementary Figures

**Sup. Fig. 1:**
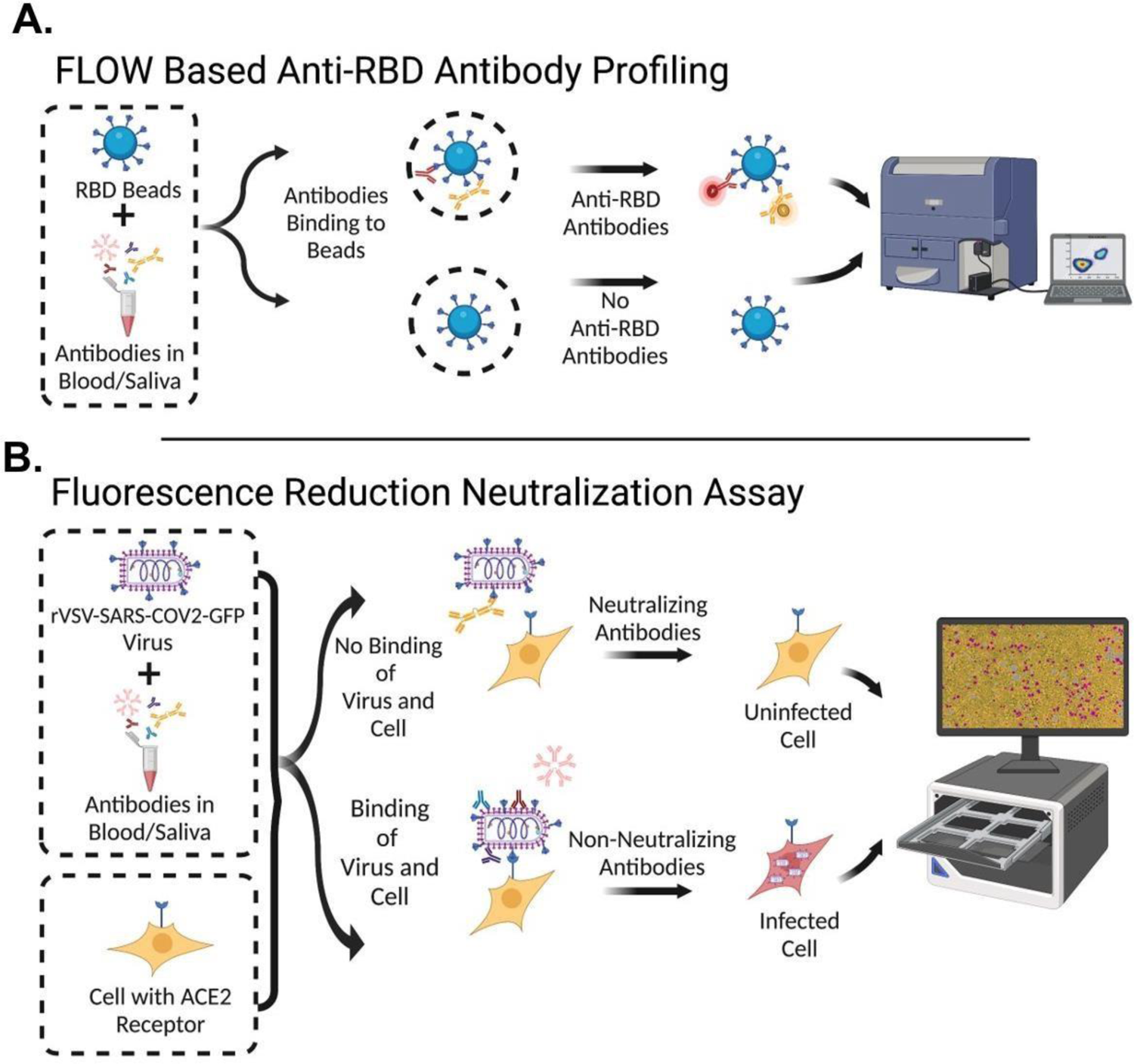
Schematic of 2 Step Flow Cytometry and FRNT Assay Workflow. **A:** Diagram of flow cytometric assay which uses RBD conjugated beads incubated with serum/saliva followed by secondary staining and analysis. **B:** Diagram of assay for measuring Fluorescence Reduction of Neutralizing Titers (FRNT) which combines pre-incubated serum/saliva and rVSV-SARS-CoV2-Spike-GFP virus with HEK293-ACE2-mCherry expressing cells. Periodic microscope imaging of cultures to quantify the amount of fluorescent GFP/mCherry signal is then performed in the IncuCyte.

**Sup. Fig. 2:**
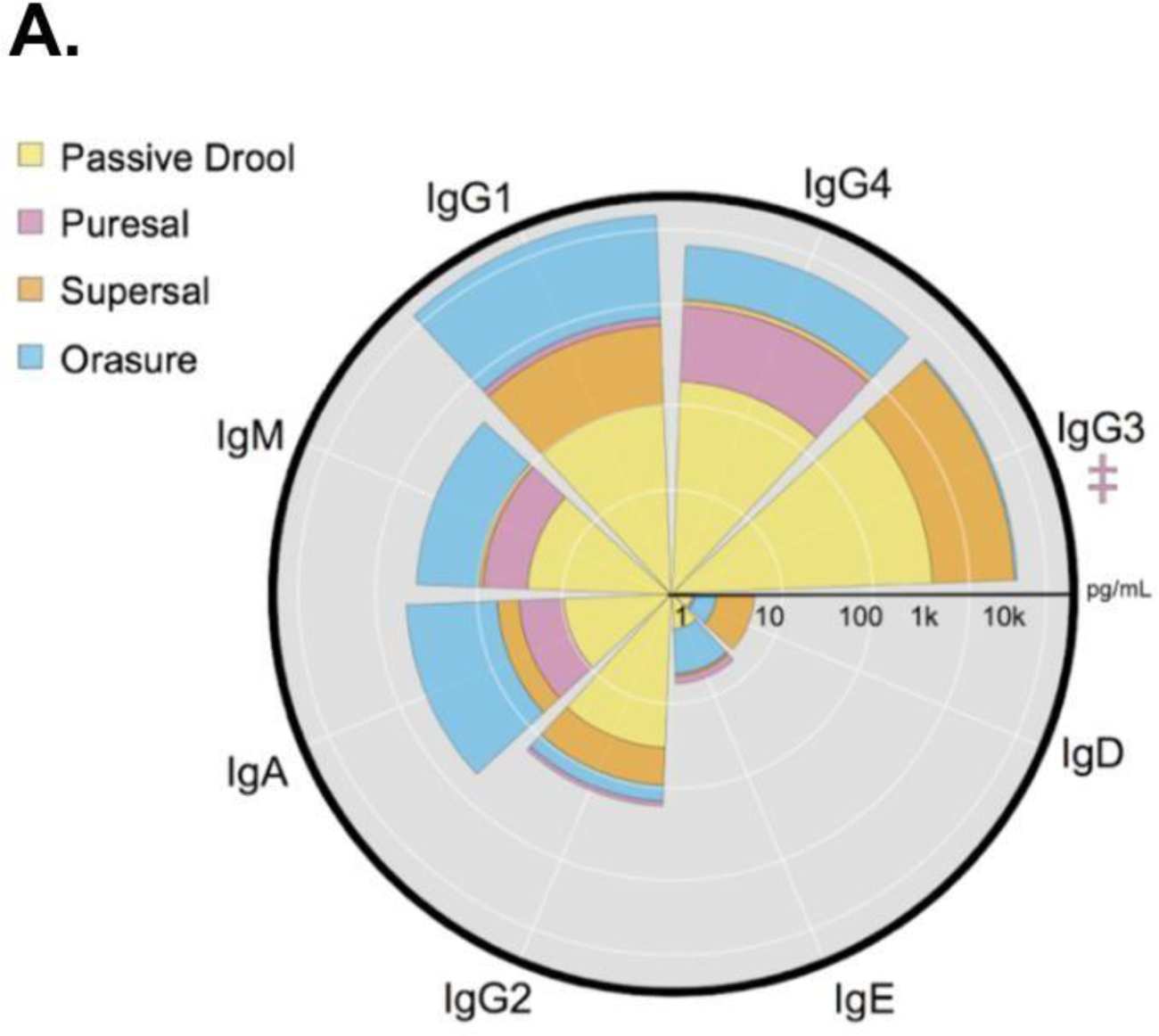
Saliva Collection Device Comparison. **A:** Comparison of 4 different saliva collection devices. The pink cross denotes the PureSal device having had an equivalent value of the OraSure (maximum value of detection for IgG3).

**Sup. Fig 3:**
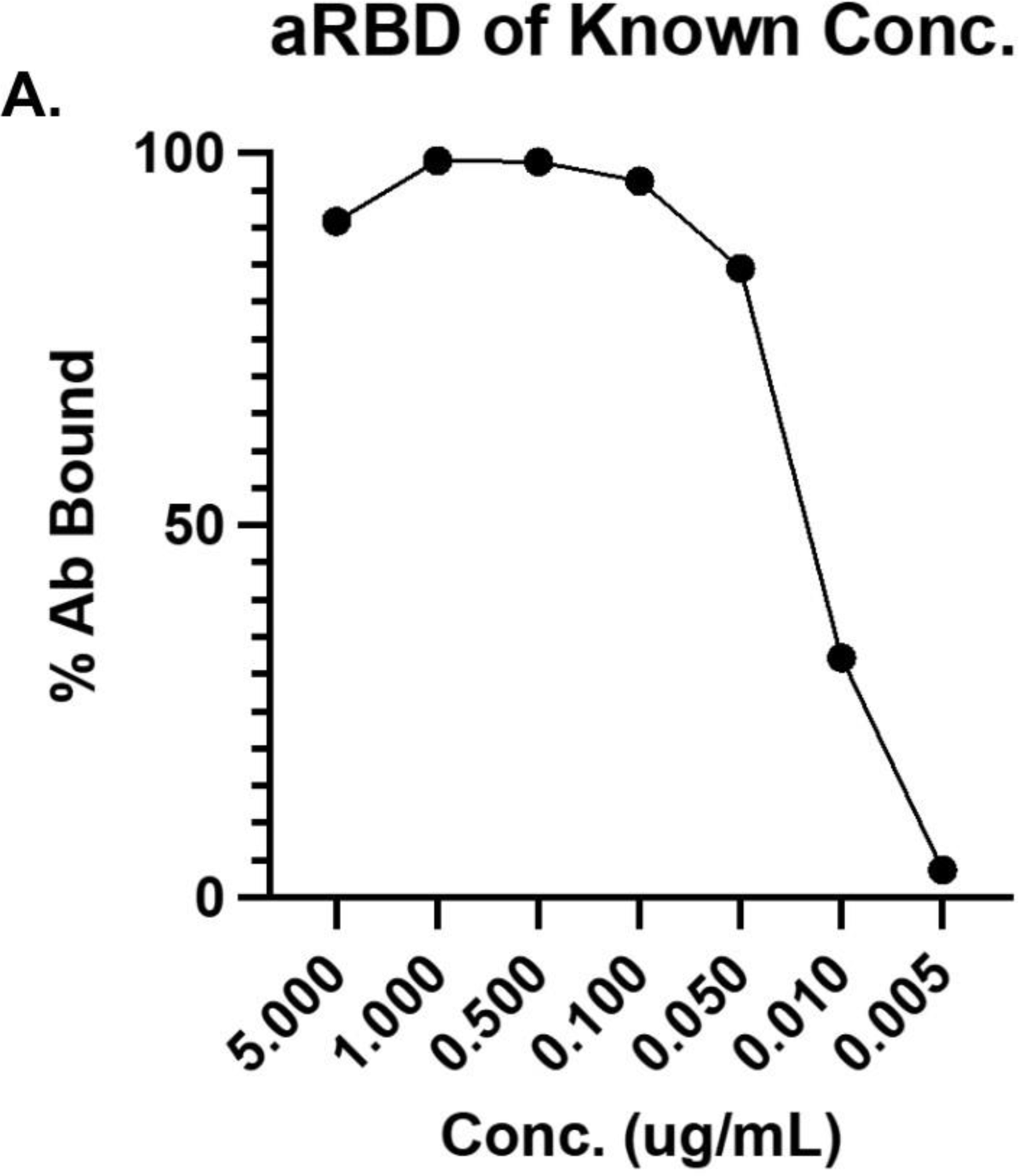
anti-RBD Binding of Known Concentration. **A:** Anti-RBD monoclonal antibody was incubated with RBD-conjugated beads at manufacturers recommended concentrations. Bead bound monoclonal anti-RBD was detected using secondary AF488 labeled IgG to evaluate the sensitivity of RBD coated BioLegend beads via flow cytometric analysis.

**Sup. Fig. 4:**
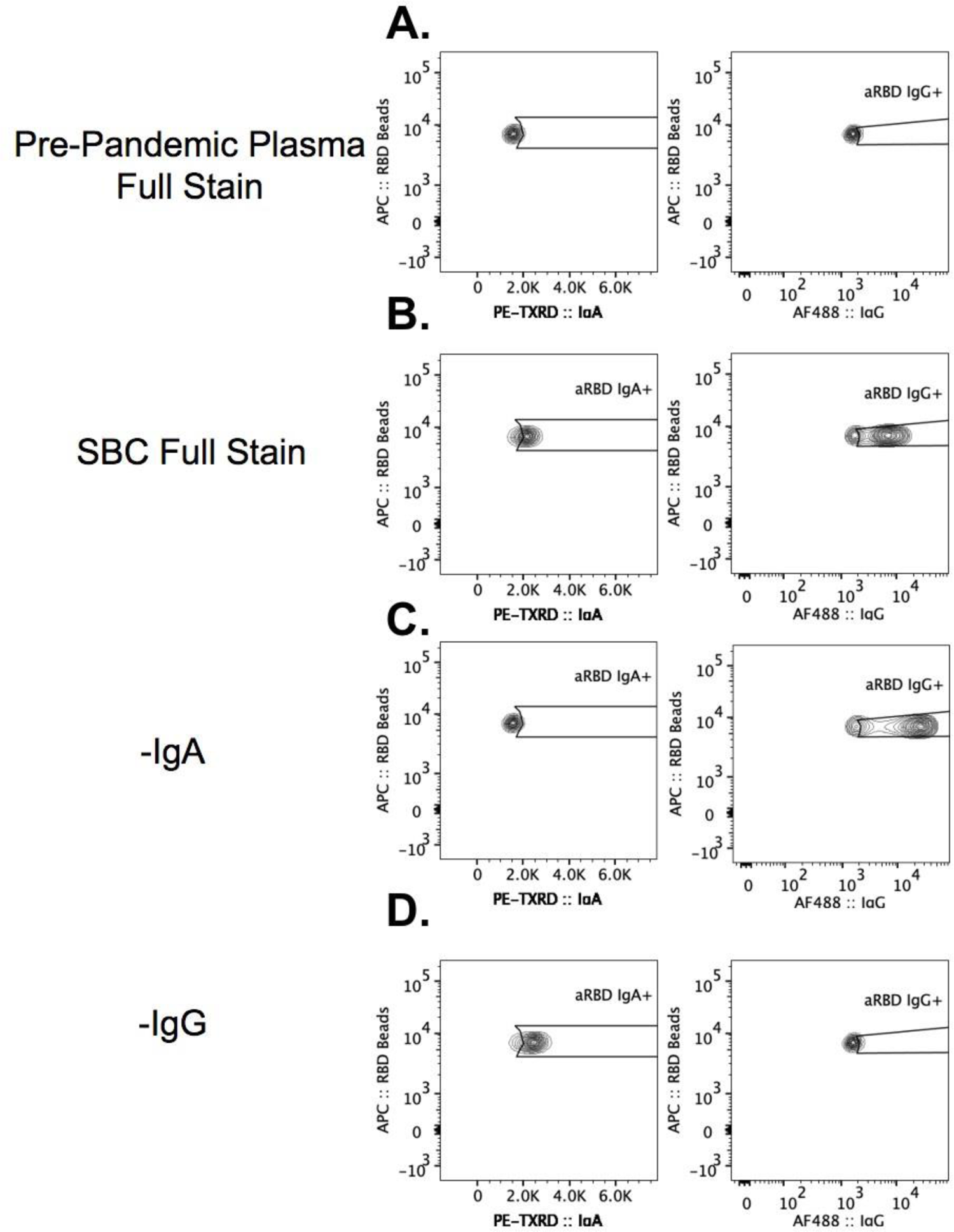
FMO in Plasma for anti-RBD IgA and IgG Fluorescence Minus One (FMO) controls of PE-TXRD labeled IgA and AF488 conjugated IgG of **A:** SARS-CoV-2 pre-pandemic plasma pooled samples with full stain, **B:** COVID-19 convalescent plasma from Stanford Blood Center (SBC) with full stain, **C:** FMO of PE-TXRD in SBC plasma, **D:** FMO of AF488 in SBC plasma.

**Sup. Fig. 5:**
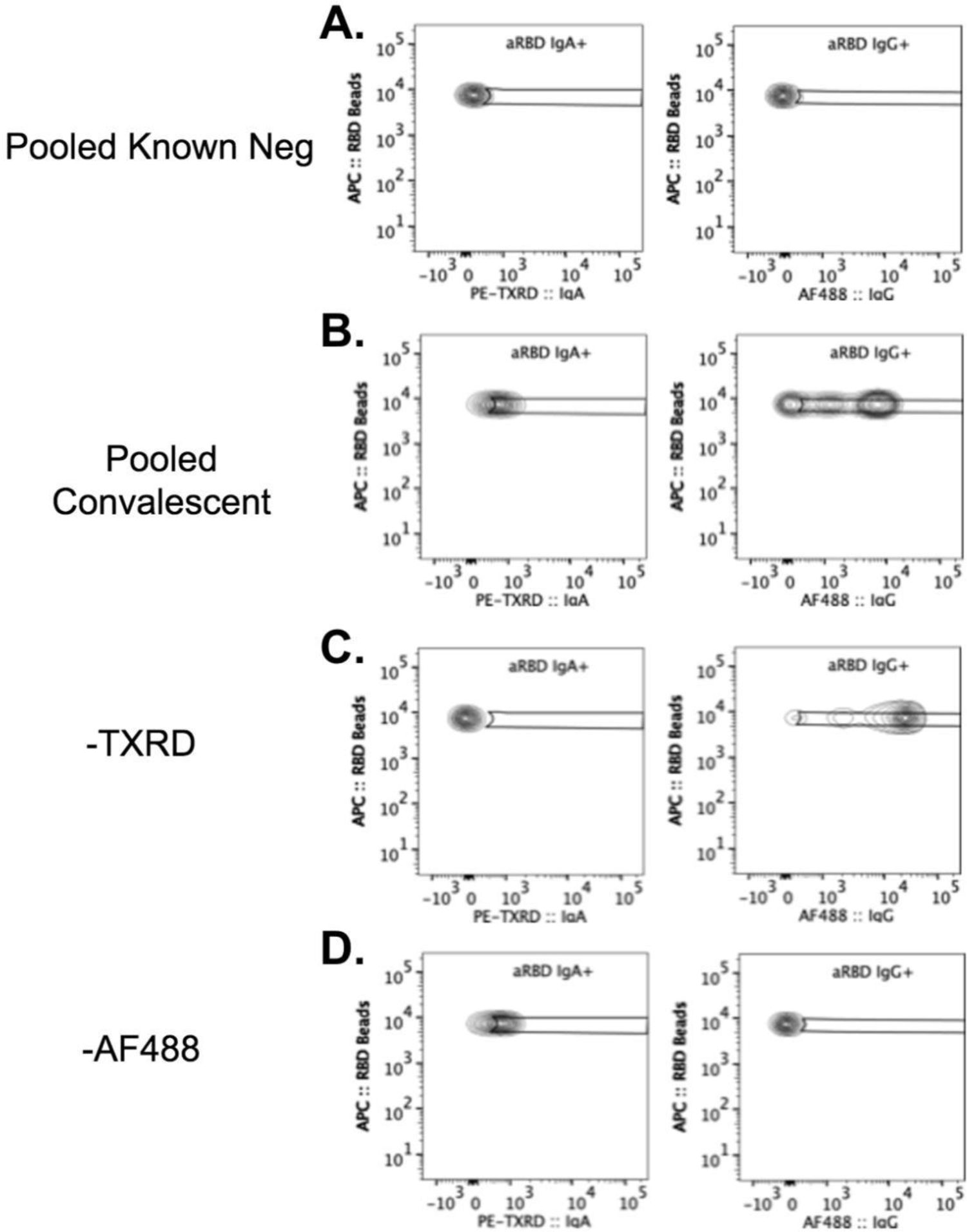
Fluorescence Minus One (FMO) in Saliva Fluorescence Minus One (FMO) controls of PE-TXRD labeled IgA and AF488 conjugated IgG of **A:** Known negative pooled saliva samples with full stain, **B:** COVID-19 convalescent saliva with full stain, **C:** FMO of PE-TXRD in convalescent saliva, **D:** FMO of AF488 in convalescent saliva.

**Sup. Fig. 6:**
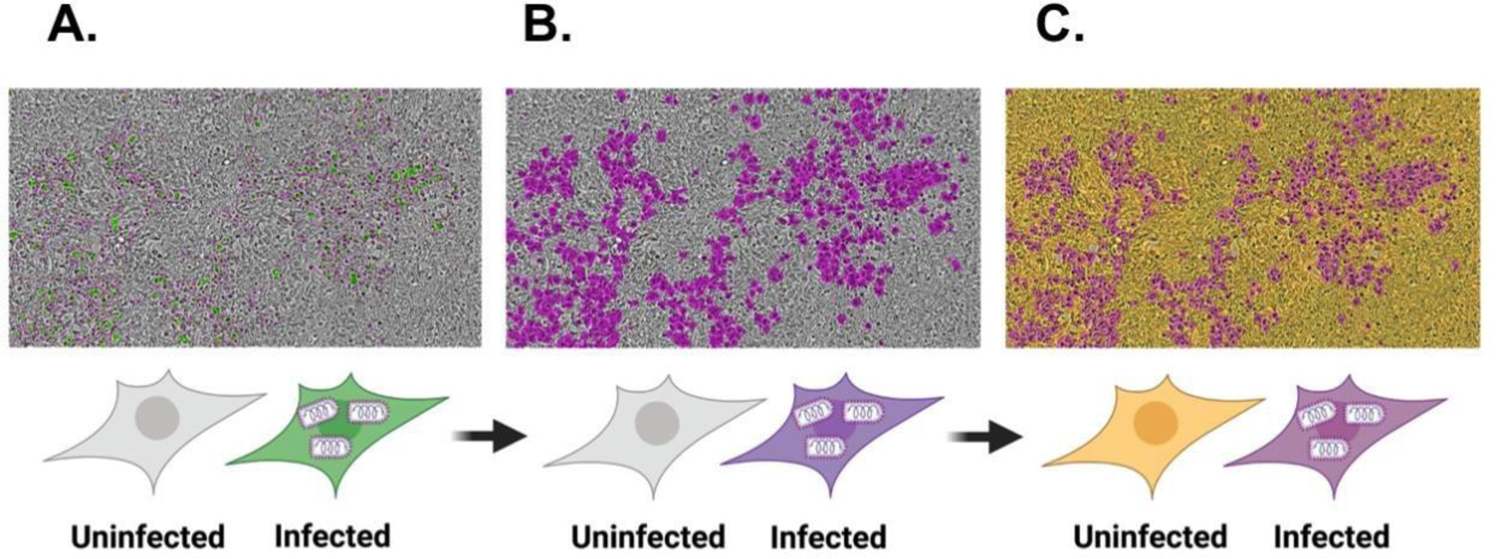
Detection of rVSV-SARS-CoV2-GFP Virus Fluorescence in GFP and Automated Quantification of Infected Cells Analysis of neutralization by IncuCyte imaging combines **A:** HEK293-ACE2-mCherry expressing cells with rVSV-SARS-COV2-GFP virus to monitor infection based on the intensity of GFP fluorescence, **B:** A purple mask created using the IncuCyte software to change the color of the infected cells from green to purple so that the infected cells presented in a colorblind friendly palette, and **C:** a yellow mask created over uninfected HEK293-ACE2-mCherry expressing cells to enhance the contrast of the neutralization visualization.

**Sup. Fig. 7:**
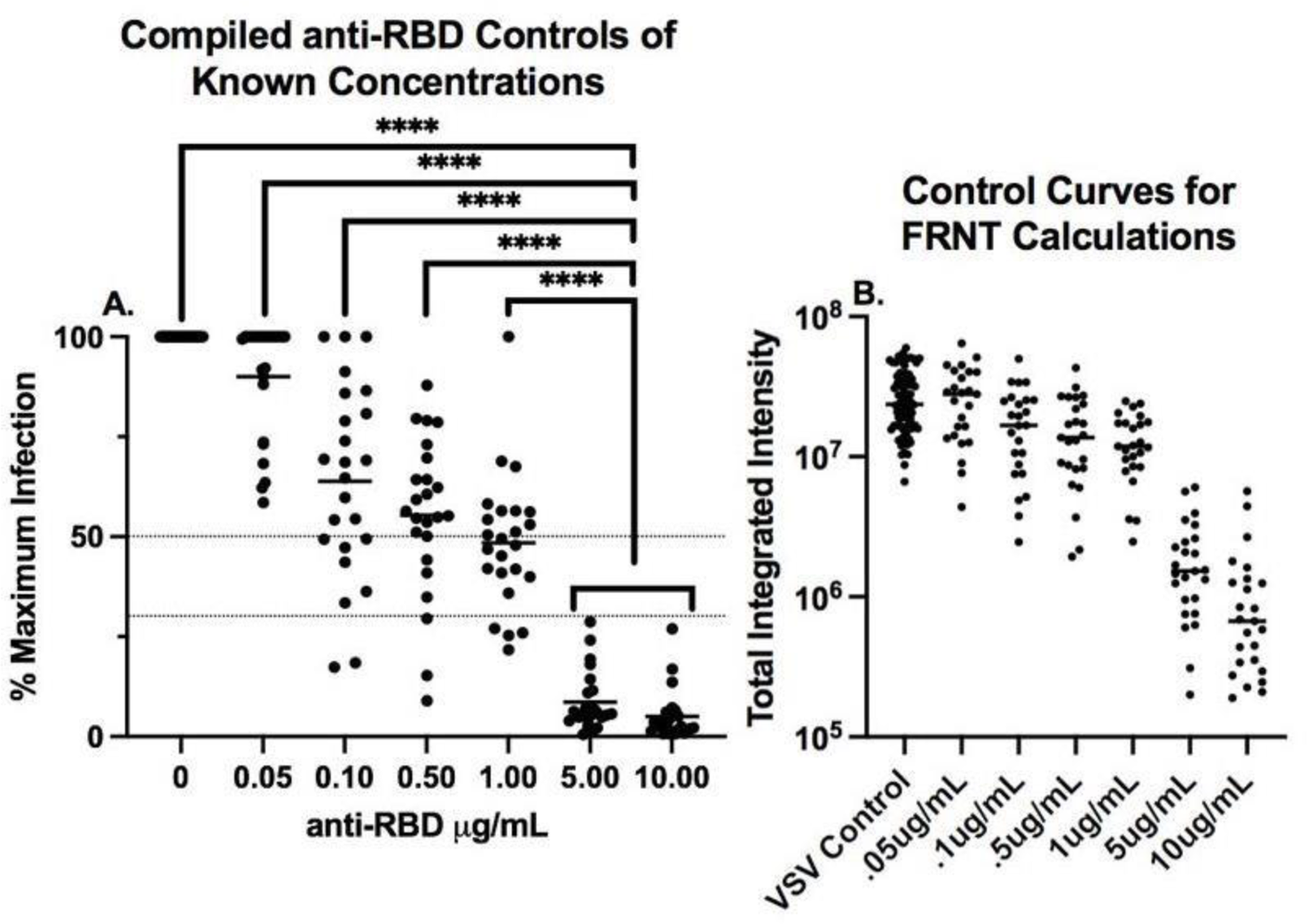
Ad26.COV2.S FRNT Assay anti-RBD Curve Control **A**: Compilation of anti-RBD monoclonal antibody curves from the neutralization assays used to determine the normalization strategy for each plate used to analyze the convalescent, Ad26.COV2.S, and mRNA samples in this study. Each dot represents the fluorescent signal at 72 hrs after adding diluted sample as indicated and VSV to the cells, displayed as percent of maximum of the triplicate rVSV-eGFP-SARS-CoV-2 control conditions or of the 0.05 μg/mL antiRBD condition. Plates were normalized to the 0.05 μg/mL anti-RBD condition only if division by the triplicate rVSV-eGFP-SARS-CoV-2 control conditions resulted in loss of a sigmoidal shape of the anti-RBD curve. Any values over 100% of maximum infection were deemed as saturated and as such a threshold of 100% was set. The distribution of points in the control samples of 5 and 10ug μg/mL anti-RBD neutralizing antibodies fit a gamma function with gamma fit parameters: alpha = 1.21638430141118, scale = 5.6113997806159. With these distribution values the chance of a point with >5ug μg/mL of neutralizing antibody being above 50 is 0.00024 and the chance of a point being above 30 is 0.00776. Fluorescence reduction in neutralization titer (FRNT) of rVSV-eGFP-SARS-CoV-2 infection of HEK293-hACE2-mCherry cells was assessed at both thresholds of normalized percentage of maximum fluorescence of 50 (FRNT50) or below and 30 or below (FRNT70) where the differences between samples with 0.05 μg/mL or less neutralizing antibodies and more than 5 μg/mL or more neutralizing antibodies in the sample are highly significant. Mann-Whitney U tests were used for comparisons (\*\*\*\**p* ≤.0001) **B**: Comparison of the raw value of the total integrated intensity of 5 μg/mL anti-RBD, 10 μg/mL anti-RBD, and VSV only control conditions from all FRNT calculations performed. Pluses denote .05 μg/mL anti-RBD conditions. Due to natural variance in total integrated intensity between plates, all plates were normalized as described in A to allow for comparison.

**Sup. Fig. 8:**
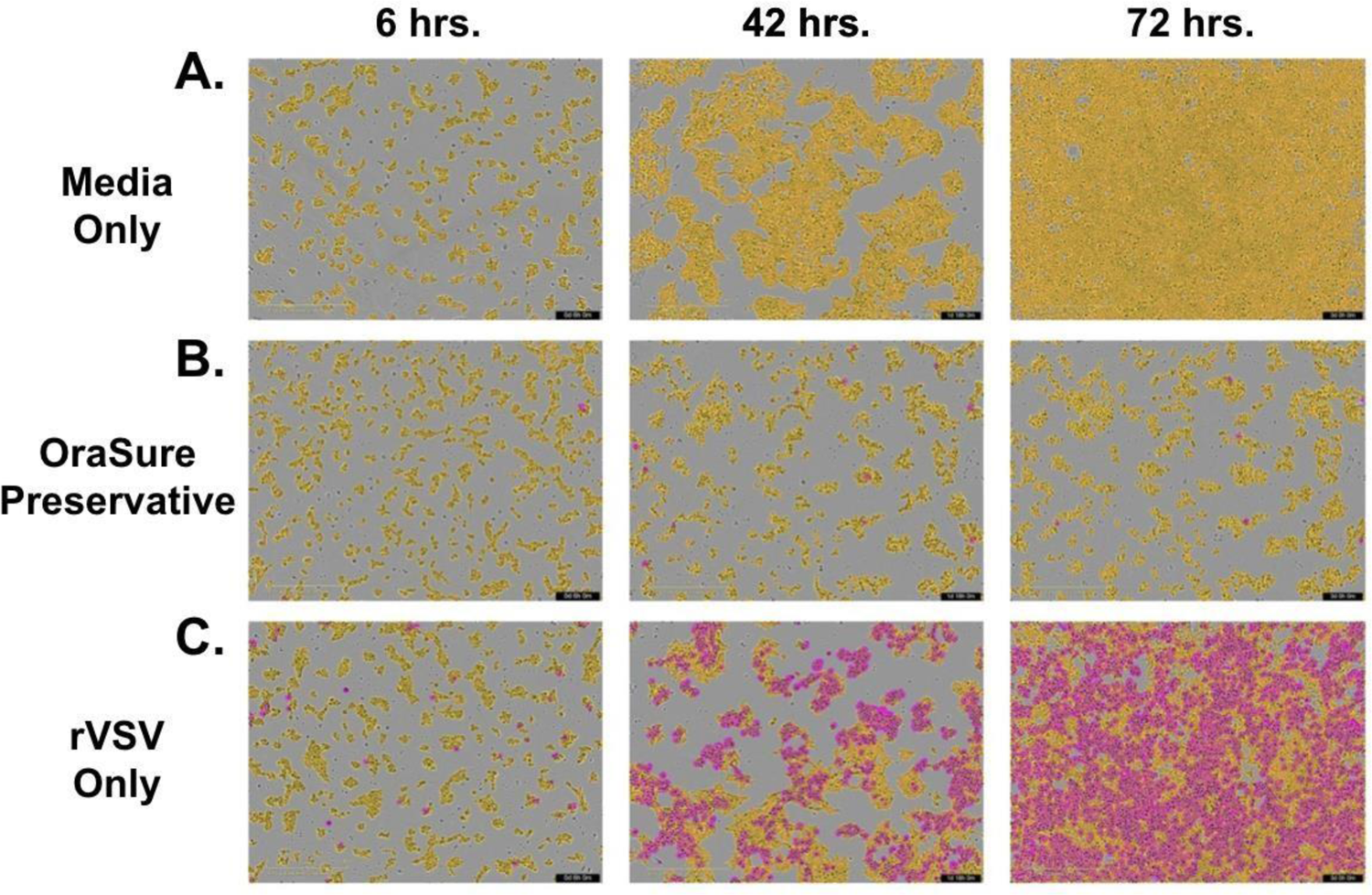
OraSure Preservative Results in Cell Toxicity at 1:15 Dilution IncuCyte images of control conditions used in the FRNT assay, collected over 3 timepoints beginning at 6 hours, 42 hours, and 72 hours including **A:** HEK293-ACE2-mCherry cells seeded at a density of 25,000 cells/well in culture media, **B:** standard OraSure preservative at a dilution of 1:15, and **C:** rVSV-eGFPSARS-CoV-2 at experimentally determined volumes for that batch with PBS supplemented in place of participant sample.

**Sup. Fig. 9:**
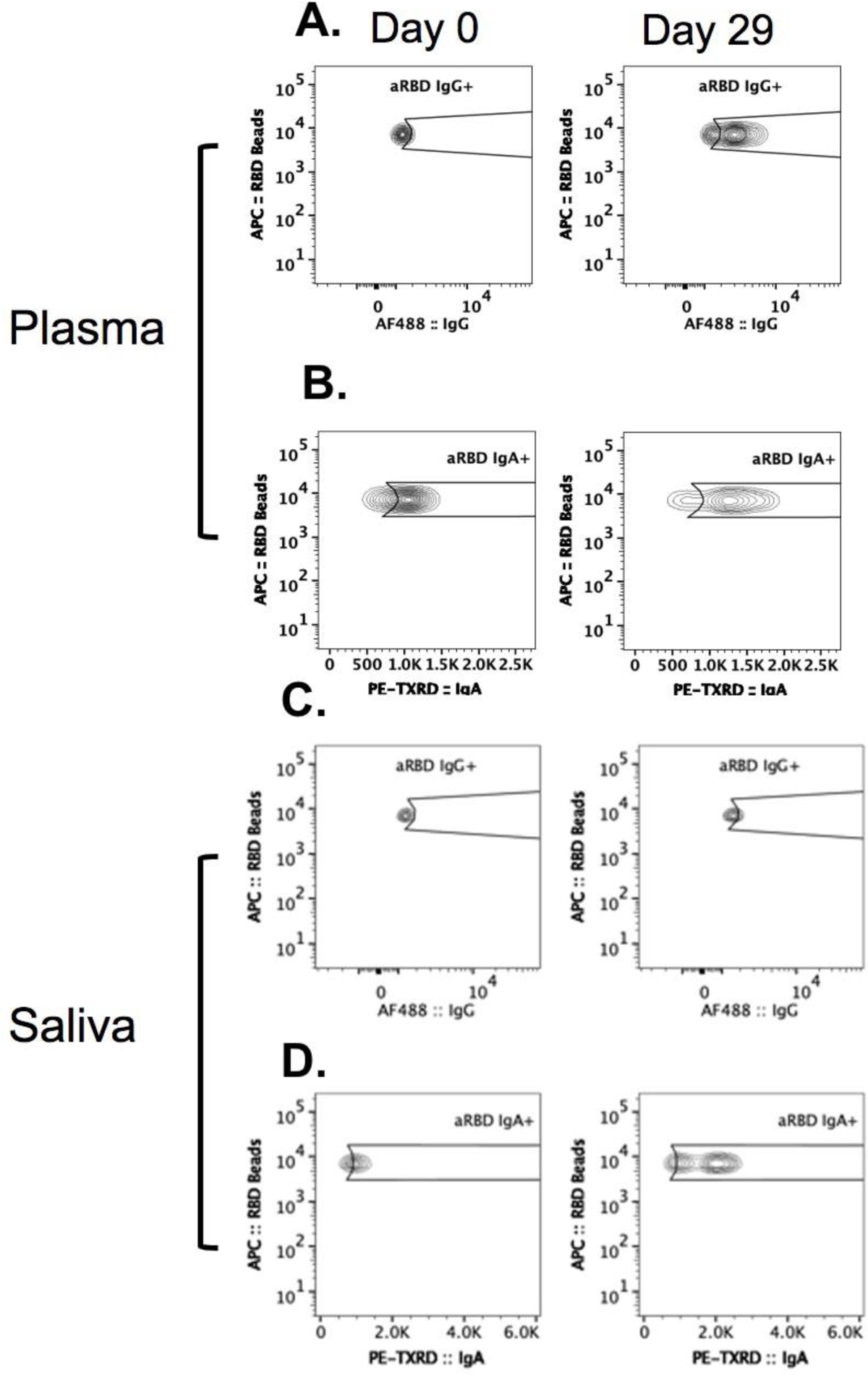
Ad26.COV2.S placebo recipient had detectable levels of anti-RBD IgG and IgA at d29 in plasma and saliva. This study participant was reported as having received the Ad26.COV2.S placebo saline injection after unblinding but was excluded from reported results herein due to detectable **A:** plasma IgG against RBD that increased from day 0 to day 29 and **B:** high levels of plasma IgA against RBD at days 0 (+/- 3 days) and 29 (+/- 3 days) by flow cytometric analysis. Further, while matched saliva samples **C:** do not show detectable anti-RBD IgG at day 0 (+/- 3 days) or day 29 (+/- 3 days), **D:** anti-RBD IgA increases from day 0 (+/- 3 days) to day 29 (+/- 3 days) by flow cytometric analysis.

**Sup. Fig. 10:**
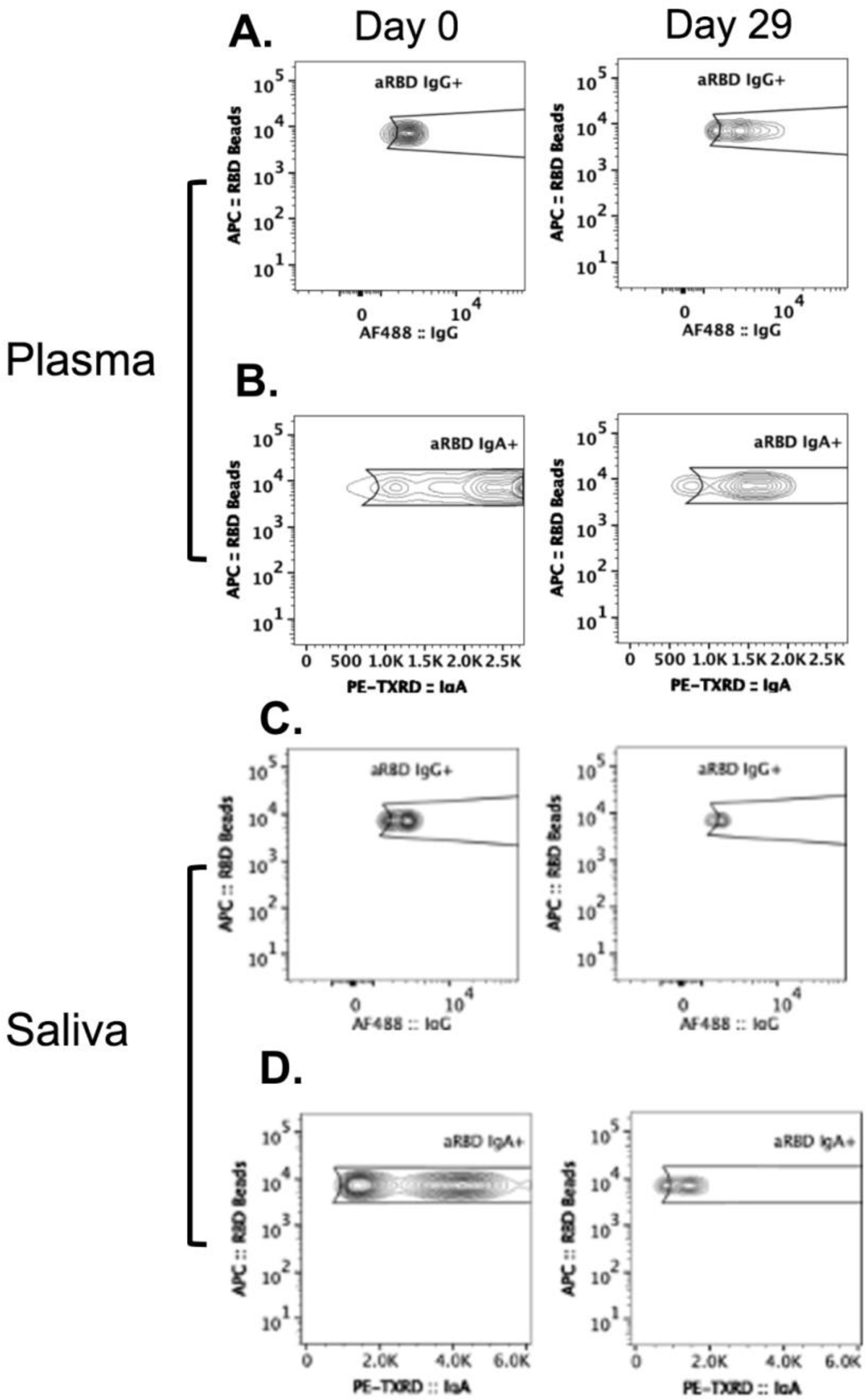
Ad26.COV2.S placebo recipient had detectable levels of anti-RBD IgG and IgA at d0 in plasma and saliva. This study participant was reported as having received the Ad26.COV2.S placebo saline injection after unblinding but was excluded from reported results herein due to detectable **A:** plasma IgG against RBD and **B:** plasma IgA against RBD at day 0 (+/- 3 days) and day 29 (+/- 3 days) by flow cytometric analysis. Matched saliva samples show detectable **C:** anti-RBD IgG and **D:** anti-RBD IgA at day 0 (+/- 3 days) and day 29 (+/- 3 days) by flow cytometric analysis.

**Sup. Fig. 11:**
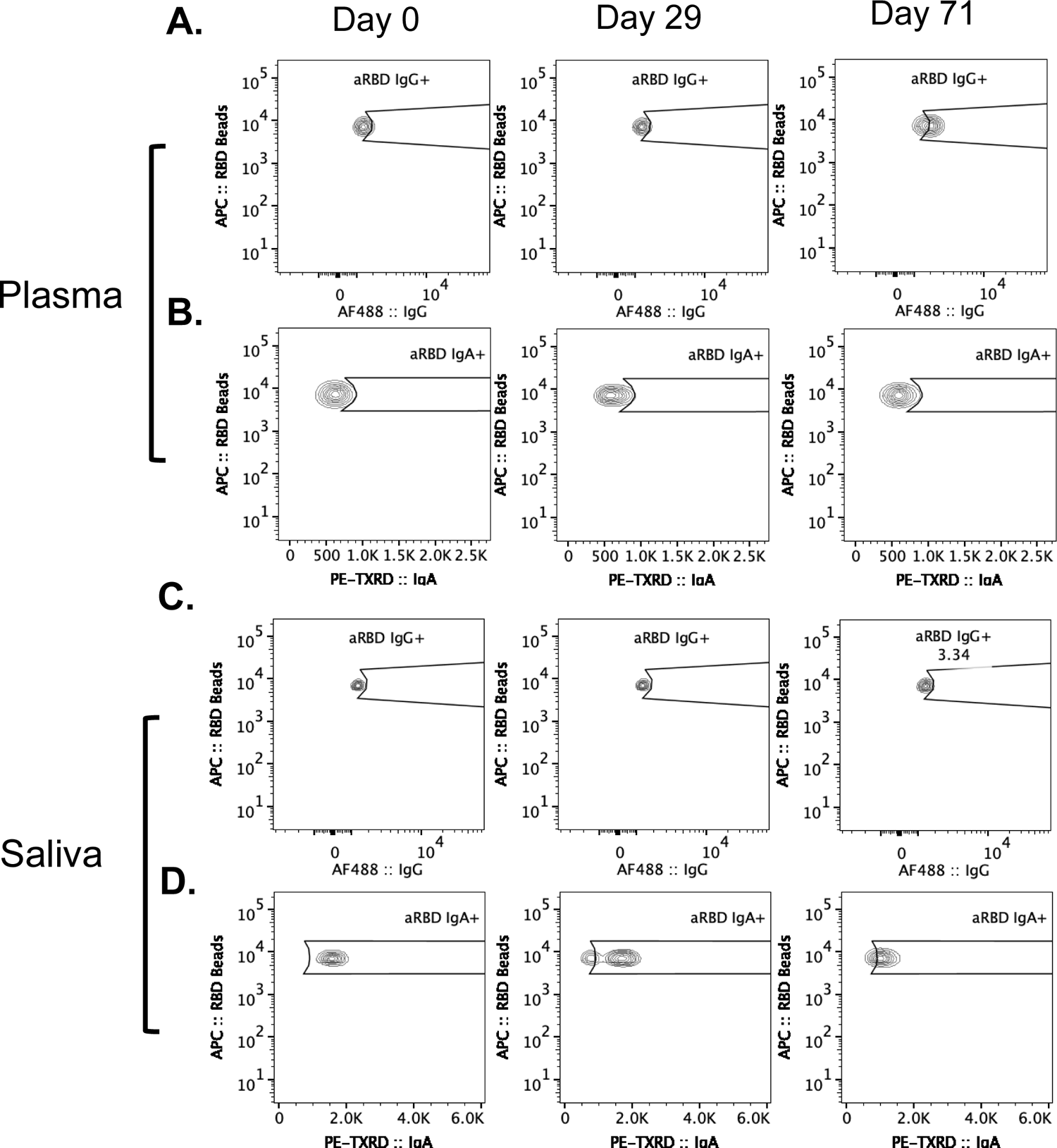
Ad26.COV2.S placebo recipient had detectable levels of salivary anti-RBD IgA at d0, d29, d71 and plasma IgG at d71. This study participant was reported as having received the Ad26.COV2.S placebo saline injection after unblinding but was excluded from reported results herein due to detectable **A:** plasma IgG against RBD at day 71 (+/- 3 days) while **B:** plasma IgA remained undetectable by flow cytometric analysis. Matched saliva samples show **C:** undetectable salivary anti-RBD IgG while **D:** salivary anti-RBD IgA is detectable at day 0, day 29 (+/- 3 days) at day 71 (+/- 3 days) by flow cytometric analysis.

**Sup. Fig. 12:**
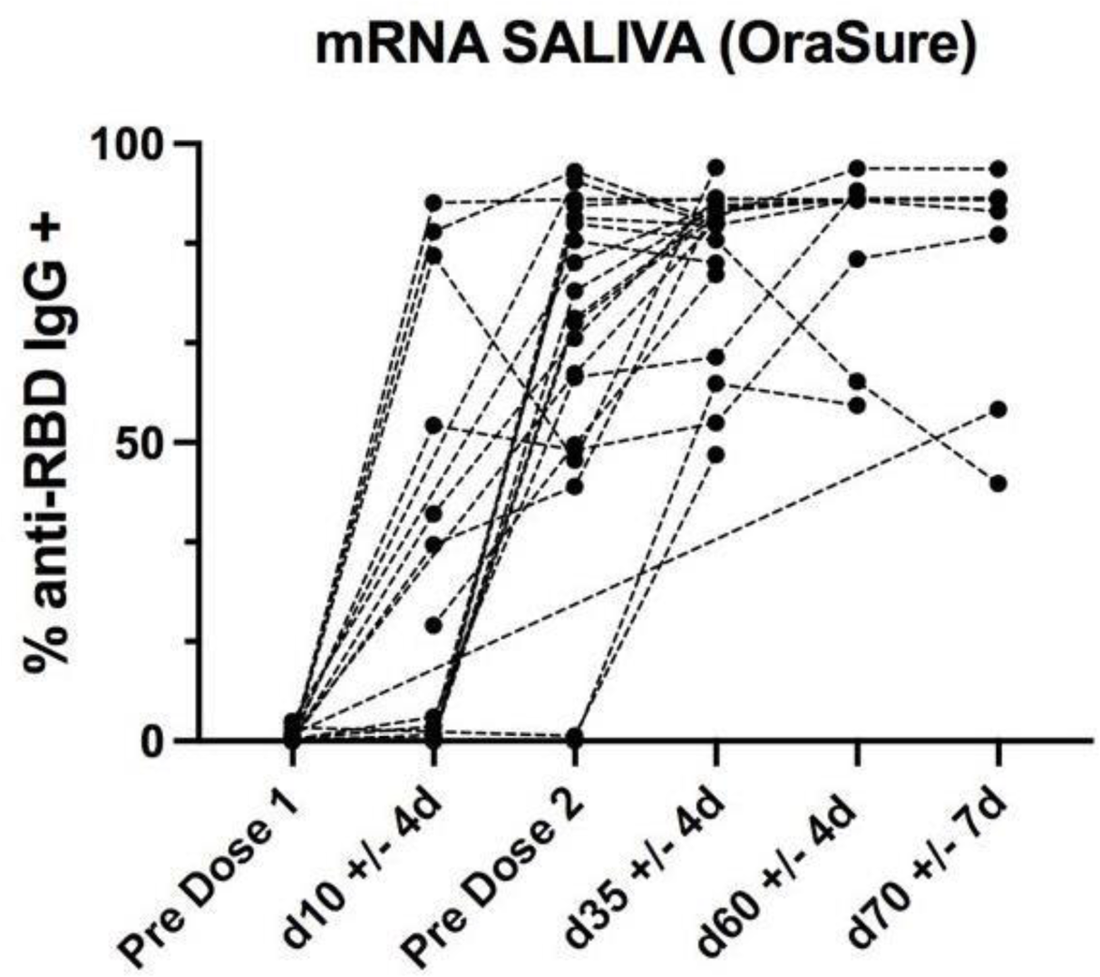
Longitudinal Saliva Sampling of 2-dose mRNA-1273/BNT162b2 Vaccine Recipients. Saliva samples collected using OraSure from study participants (n=23) who received 2 doses of either BNT162b2 or mRNA-1273 mRNA vaccines. Samples were collected prior to receiving the vaccine dose on the same day as the first or second vaccine dose was received (Pre Dose 1 and Pre Dose 2 respectively). Subsequent samples were collected according to the different vaccine schedules and then after both doses on day 35 +/- 4 days, at day 60 +/- 4 days and at day 70 +/- 7 days. Across participants, some sample time-points were not collected and therefore not shown in this data.

**Sup. Fig. 13:**
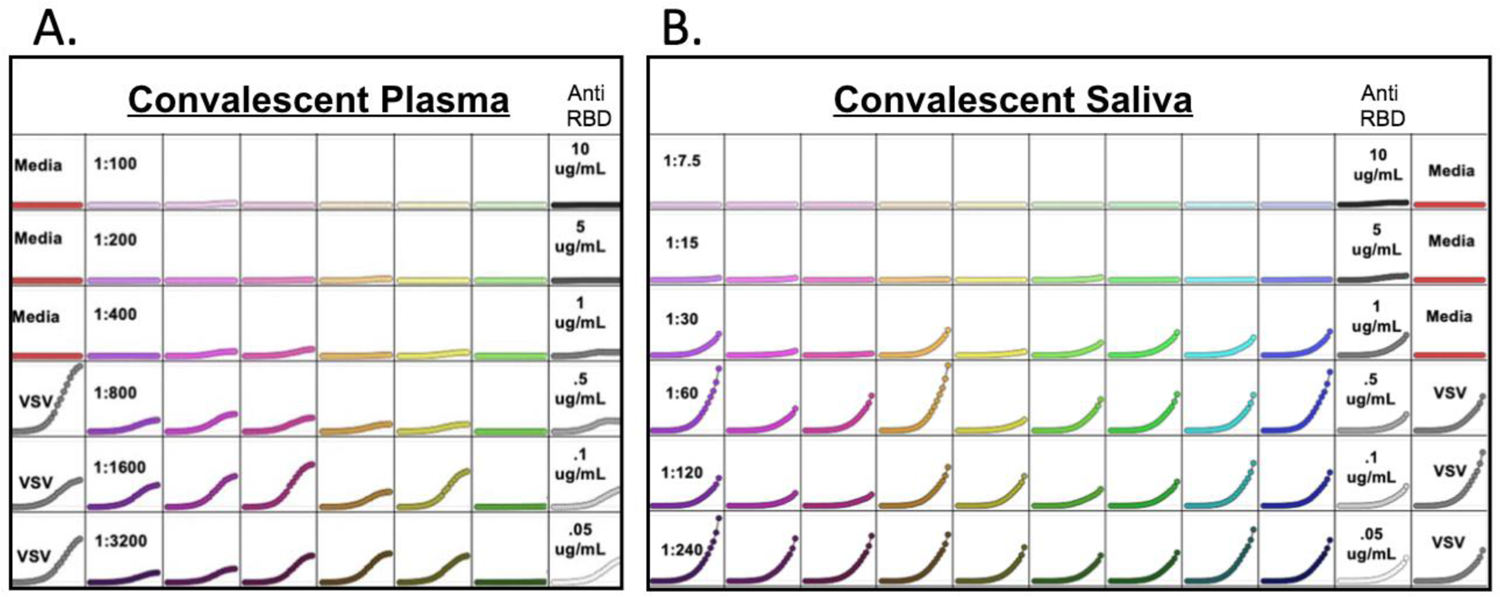
Raw data of Fluorescent Reduction Neutralization Assay from Convalescent Plasma and Saliva Representative plates of total integrated intensity of GFP Fluorescence over 72 hours from Fluorescence Reduction Neutralization Assay of **A:** convalescent plasma and **B:** convalescent saliva. Each plate contains an anti-RBD monoclonal dilution series as well as triplicate control wells of rVSV-SARS-COV2-GFP and HEK293-ACE2-mCherry media

**Sup. Fig. 14:**
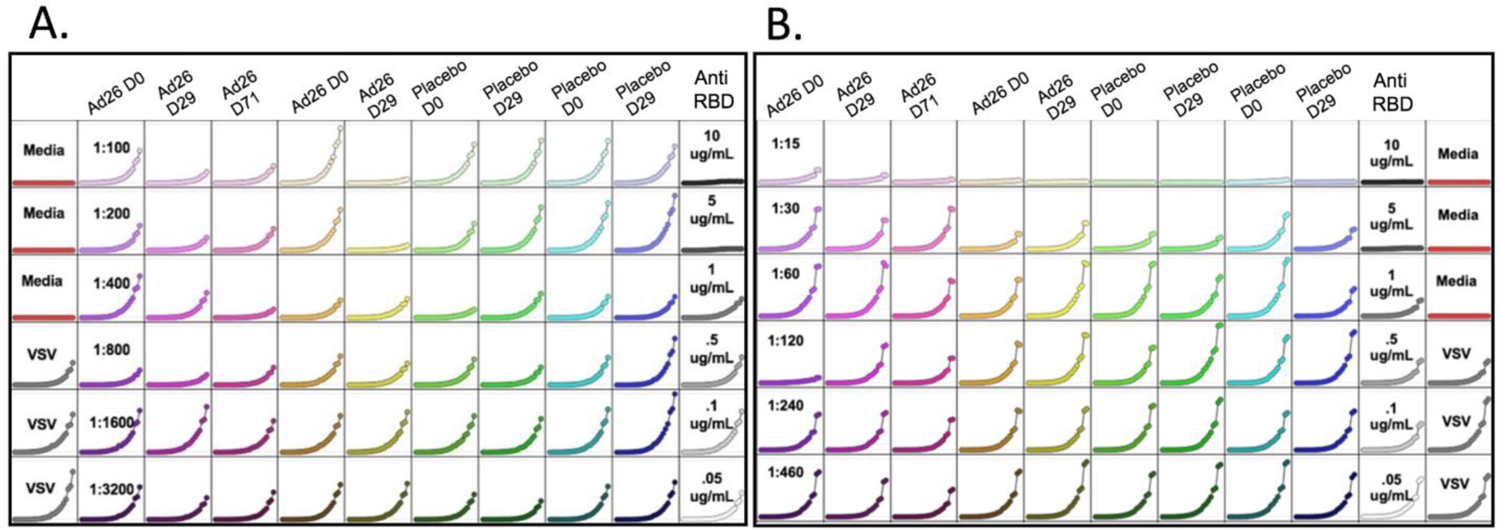
Raw data of Fluorescent Reduction Neutralization Assay from Ad26.COV2.S Plasma and Saliva Representative plates of total integrated intensity of GFP Fluorescence over 72 hours from Fluorescence Reduction Neutralization Assay of **A:** Ad26.COV2.S plasma and **B:** AD26.COV2.S saliva. Each plate contains an anti-RBD monoclonal dilution series as well as triplicate control wells of rVSV-SARS-COV2-GFP and HEK293-ACE2-mCherry media

**Sup. Fig. 15:**
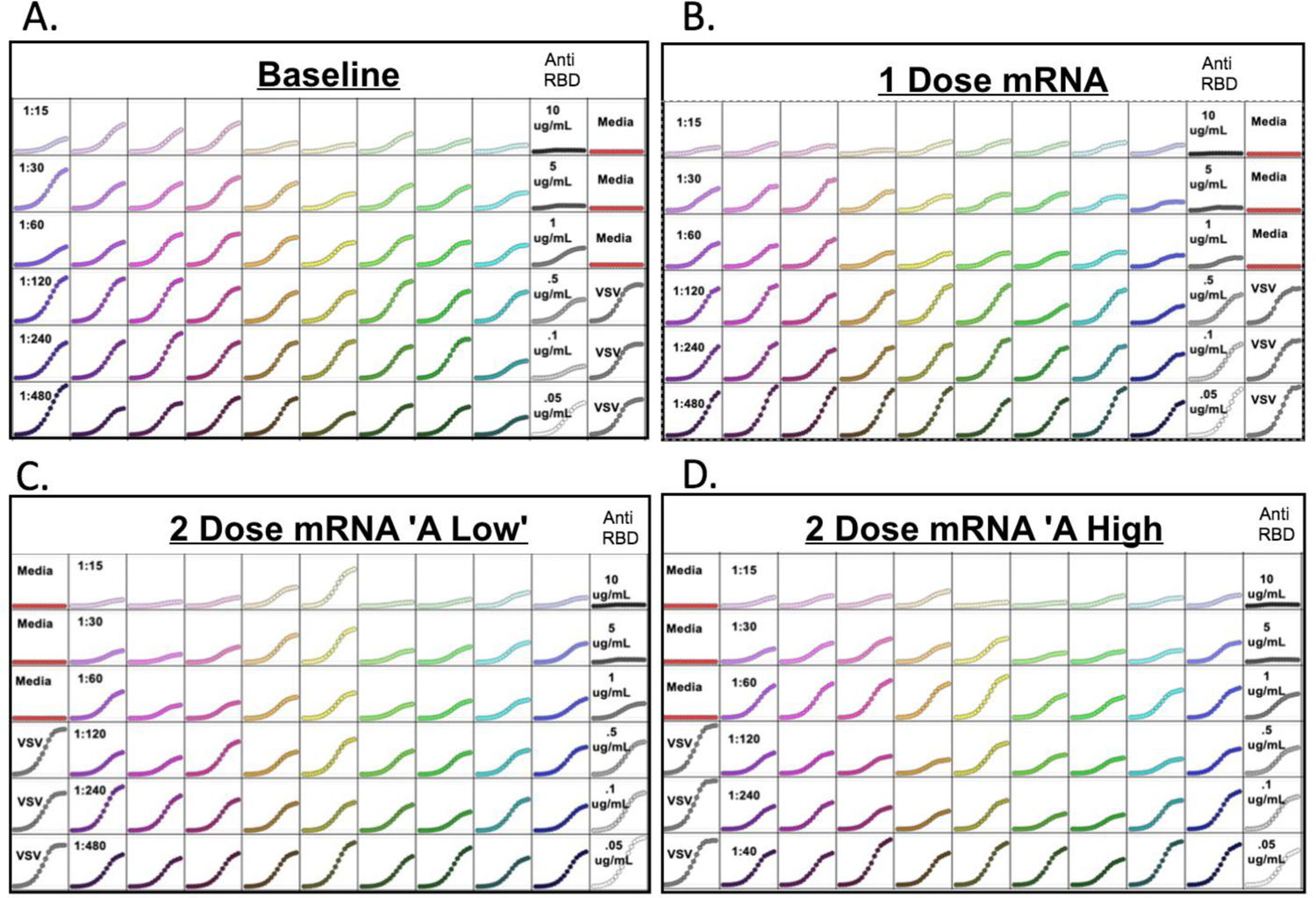
Raw data of Fluorescent Reduction Neutralization Assay from mRNA Saliva. Representative plates of total integrated intensity of GFP Fluorescence over 72 hours from Fluorescence Reduction Neutralization Assay of **A:** baseline saliva **B:** saliva following a single dose of an mRNA vaccine, **C-D:** saliva after 2 doses of an mRNA vaccine designated as **C:** A low or **D:** A high as designated by the Gommerman Lab following an ELISA assay. Each plate contains an antiRBD monoclonal dilution series as well as triplicate control wells of rVSV-SARS-COV2-GFP and HEK293-ACE2-mCherry media

## Supplementary Tables

**Sup. Table 1:** Ad26.COV2.S Vaccine Study Participants

| IRB 57277 Samples: |  |  |  |
| --- | --- | --- | --- |
| Subject Age | Biological Sex Assigned at Birth | Sample Type | Sampling Time Points |
| 70-79 | M | Ad26.COV2.S | Day 0, 29 |
| 60-69 | F | Ad26.COV2.S | Day 0, 29, 71 |
| 50-59 | F | Ad26.COV2.S | Day 0, 29 |
| 40-49 | M | Ad26.COV2.S | Day 0, 29 |
| 60-69 | F | Ad26.COV2.S | Day 0 |
| 80-90 | F | Ad26.COV2.S | Day 0, 29 |
| 60-69 | F | Ad26.COV2.S | Day 0, 29 |
| 60-69 | M | Ad26.COV2.S | Day 0, 29, 71 |
| 80-89 | F | Ad26.COV2.S | Day 0 |
| 60-69 | M | Ad26.COV2.S | Day 0, 29, 71 |
| 60-69 | M | Ad26.COV2.S | Day 0, 29, 71 |
| 60-69 | F | Ad26.COV2.S | Day 0, 29 |
| 40-49 | F | Ad26.COV2.S | Day 0, 29, 71 |
| 60-69 | M | Ad26.COV2.S | Day 0, 29 |
| 40-49 | F | Ad26.COV2.S | Day 0, 29 |
| 60-69 | M | Ad26.COV2.S | Day 0, 29 |
| 40-49 | F | Ad26.COV2.S | Day 0, 29, 71 |
| 40-49 | F | Ad26.COV2.S | Day 0, 29, 71 |
| 50-59 | F | Ad26.COV2.S | Day 0, 29 |
| 60-69 | F | Ad26.COV2.S | Day 0, 29, 71 |
| 50-59 | F | Ad26.COV2.S | Day 0, 29 |
| 50-59 | F | Ad26.COV2.S | Day 0, 29, 71 |
| 50-59 | M | Ad26.COV2.S | Day 0, 29 |
| 50-59 | F | Ad26.COV2.S | Day 0, 29, 71 |
| 30-39 | M | Ad26.COV2.S | Day 0, 29, 71 |
| 30-39 | M | Ad26.COV2.S | Day 0, 29 |
| 50-59 | M | Ad26.COV2.S | Day 0, 29 |
| 20-29 | F | Ad26.COV2.S | Day 0, 29, 71 |
| 50-59 | M | Ad26.COV2.S | Day 0, 29, 71 |
| 30-39 | M | Ad26.COV2.S | Day 0, 29, 71 |
| 40-49 | F | Ad26.COV2.S | Day 0, 29 |
| 20-29 | F | Ad26.COV2.S | Day 0, 29 |
| 20-29 | M | Ad26.COV2.S | Day 0, 29, 71 |
| 50-59 | M | Placebo | Day 0, 29 |
| 70-79 | M | Placebo | Day 0, 29 |
| 60-69 | F | Placebo | Day 0, 29 |
| 40-49 | F | Placebo | Day 0, 29 |
| 60-69 | F | Placebo | Day 0, 29, 71 |
| 40-49 | M | Placebo | Day 0, 29 |
| 60-69 | M | Placebo | Day 0, 29 |
| 70-79 | F | Placebo | Day 0, 29 |
| 60-69 | M | Placebo | Day 0, 29, 71 |
| 60-69 | M | Placebo | Day 0, 29, 71 |
| 60-69 | M | Placebo | Day 0, 29 |
| 60-69 | F | Placebo | Day 0, 29, 71 |
| 50-59 | M | Placebo | Day 0, 29 |
| 50-59 | M | Placebo | Day 0, 29 |
| 60-69 | M | Placebo | Day 0, 29 |
| 60-69 | F | Placebo | Day 0, 29 |
| 50-59 | M | Placebo | Day 0, 29, 71 |
| 50-59 | F | Placebo | Day 0, 29 |
| 40-49 | M | Placebo | Day 0, 29, 71 |
| 70-79 | F | Placebo | Day 0, 29 |
| 60-69 | M | Placebo | Day 0, 29, 71 |
| 30-39 | M | Placebo | Day 0, 29, 71 |
| 60-69 | M | Placebo | Day 0, 29 |
| 40-49 | M | Placebo | Day 0, 29 |
| 40-49 | M | Placebo | Day 0, 29 |
| 50-59 | M | Placebo | Day 0, 29, 71 |
| 40-49 | M | Placebo | Day 0, 29 |
| 20-29 | M | Placebo | Day 0, 29, 71 |
| 50-59 | M | Placebo | Day 0, 29, 71 |
| 60-69 | M | Placebo | Day 0, 29 |
| 50-59 | F | Placebo | Day 0, 29, 71 |

**Sup. Table 2:** University of Toronto Saliva Samples

| REB 23901 Samples: |  |  |
| --- | --- | --- |
| Subject Age | Biological Sex Assigned at Birth | Sample Type |
| 60-69 | F | mRNA-1273 |
| * | * | * |
| 40-49 | F | BNT-162b2 |
| 50-59 | F | BNT-162b2 |
| 60-69 | F | mRNA-1273 |
| 40-49 | F | BNT-162b2 |
| 40-49 | F | BNT-162b2 |
| 60-69 | F | mRNA-1273 |
| 50-59 | F | BNT-162b2 |
| 50-59 | F | mRNA-1273 |
| 30-39 | M | BNT-162b2 |
| 60-69 | F | BNT-162b2 |
| 30-39 | M | BNT-162b2 |
| 30-39 | M | mRNA-1273 |
| 50-59 | F | mRNA-1273 |
| 40-49 | M | mRNA-1273 |
| 20-29 | F | mRNA-1273 |
| 50-59 | F | mRNA-1273 |
| 30-39 | F | mRNA-1273 |
| 50-59 | M | mRNA-1273 |
| 50-59 | F | mRNA-1273 |
| 50-59 | F | mRNA-1273 |
| 30-39 | F | BNT-162b2 |
| 60-69 | F | BNT-162b2 |
| 20-29 | F | BNT-162b2 |
| 60-69 | M | BNT-162b2 |
| 40-49 | F | BNT-162b2 |
| 50-59 | F | BNT-162b2 |
| 50-59 | F | BNT-162b2 |
| 100+ | * | BNT-162b2 |
| 50-59 | F | BNT-162b2 |
| 50-59 | F | BNT-162b2 |
| 20-29 | F | BNT-162b2 |
| 20-29 | F | BNT-162b2 |
| 20-29 | M | BNT-162b2 |
| 20-29 | M | BNT-162b2 |
| 20-29 | M | BNT-162b2 |
| 40-49 | F | BNT-162b2 |
| 20-29 | M | BNT-162b2 |
| 20-29 | F | BNT-162b2 |
| 30-39 | F | BNT-162b2 |
| 30-39 | F | BNT-162b2 |
| 50-59 | M | BNT-162b2 |
| 50-59 | M | BNT-162b2 |
| 20-29 | F | BNT-162b2 |
| 30-39 | M | BNT-162b2 |
| 20-29 | F | BNT-162b2 |
| 20-29 | F | BNT-162b2 |
| 30-39 | M | BNT-162b2 |
| 20-29 | M | BNT-162b2 |
| 20-29 | F | BNT-162b2 |
| 20-29 | F | BNT-162b2 |
| 20-29 | F | BNT-162b2 |
| 20-29 | F | BNT-162b2 |
| 20-29 | M | BNT-162b2 |
| 30-39 | M | BNT-162b2 |
| 40-49 | F | BNT-162b2 |
| 40-49 | F | BNT-162b2 |
| 100+ | F | mRNA-1273 |
| 20-29 | F | BNT-162b2 |
| 40-49 | M | mRNA-1273 |
| 30-39 | M | mRNA-1273 |
| 20-29 | F | BNT-162b2 |
| 20-29 | F | mRNA-1273 |
| 20-29 | F | BNT-162b2 |
| 40-49 | F | BNT-162b2 |
| 60-69 | F | mRNA-1273 |
| 60-69 | F | mRNA-1273 |
| 40-49 | F | BNT-162b2 |
| 20-29 | F | BNT-162b2 |
| 20-29 | F | mRNA-1273 |
| 60-69 | F | mRNA-1273 |
| 20-29 | F | mRNA-1273 |
| 20-29 | F | mRNA-1273 |

**Sup. Table 3:** COVID-19 Convalescent Plasma from Stanford Blood Center Samples

| IRB 57277 Samples: |  |  |
| --- | --- | --- |
| Subject Age | Biological Sex Assigned at Birth | Sample Type |
| 40-49 | F | Stanford Blood Center |
| 50-59 | F | Stanford Blood Center |
| 50-59 | M | Stanford Blood Center |
| 40-49 | M | Stanford Blood Center |
| 50-59 | F | Stanford Blood Center |
| 50-59 | M | Stanford Blood Center |
| 30-39 | M | Stanford Blood Center |
| 30-39 | M | Stanford Blood Center |
| 30-39 | F | Stanford Blood Center |
| 60-69 | M | Stanford Blood Center |
| 40-49 | M | Stanford Blood Center |
| 40-49 | F | Stanford Blood Center |
| 60-69 | M | Stanford Blood Center |
| 20-29 | F | Stanford Blood Center |
| 30-39 | F | Stanford Blood Center |
| 50-59 | F | Stanford Blood Center |
| 50-59 | F | Stanford Blood Center |
| 50-59 | M | Stanford Blood Center |
| 50-59 | M | Stanford Blood Center |
| 30-39 | M | Stanford Blood Center |

**Sup. Table 4:**
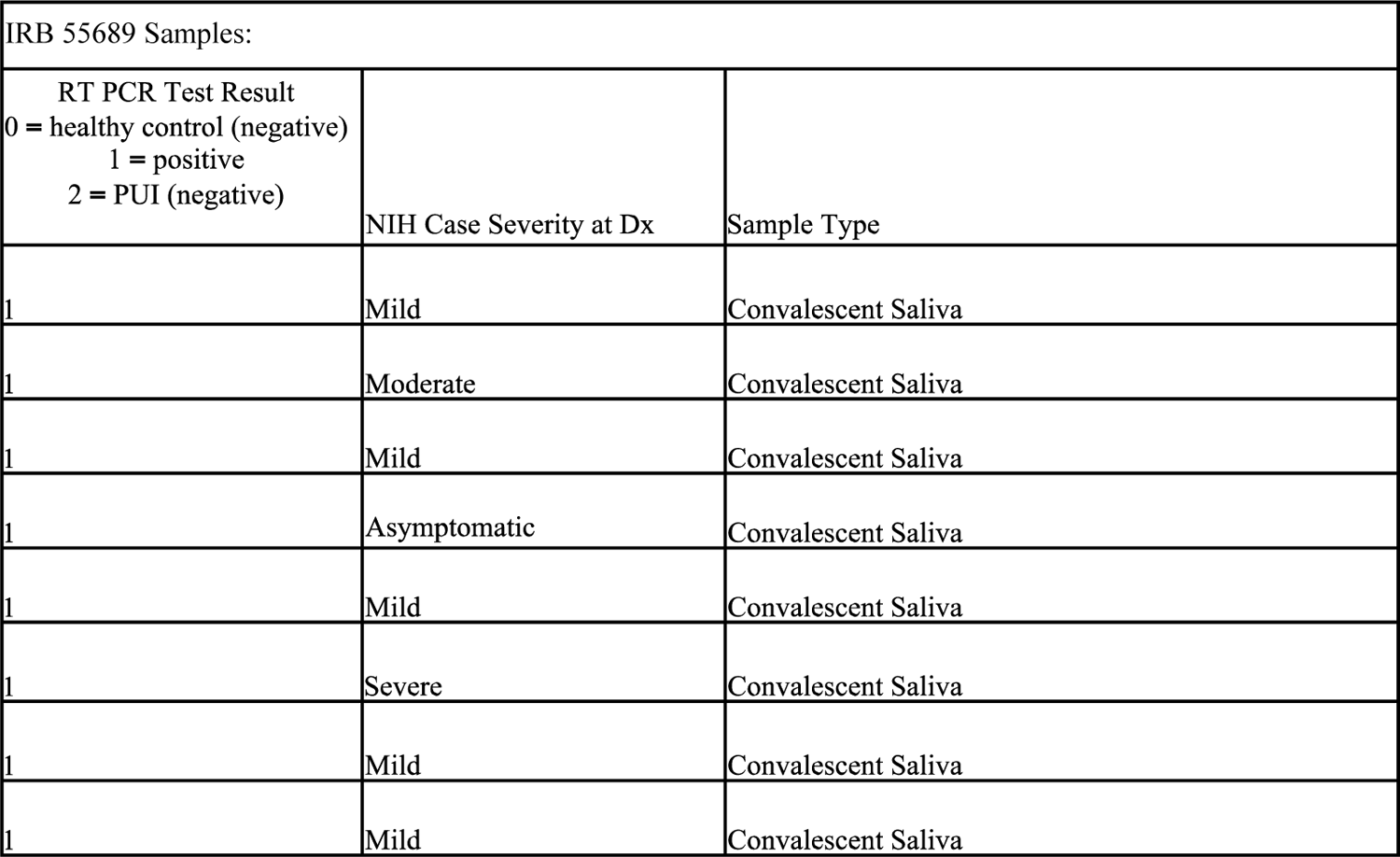
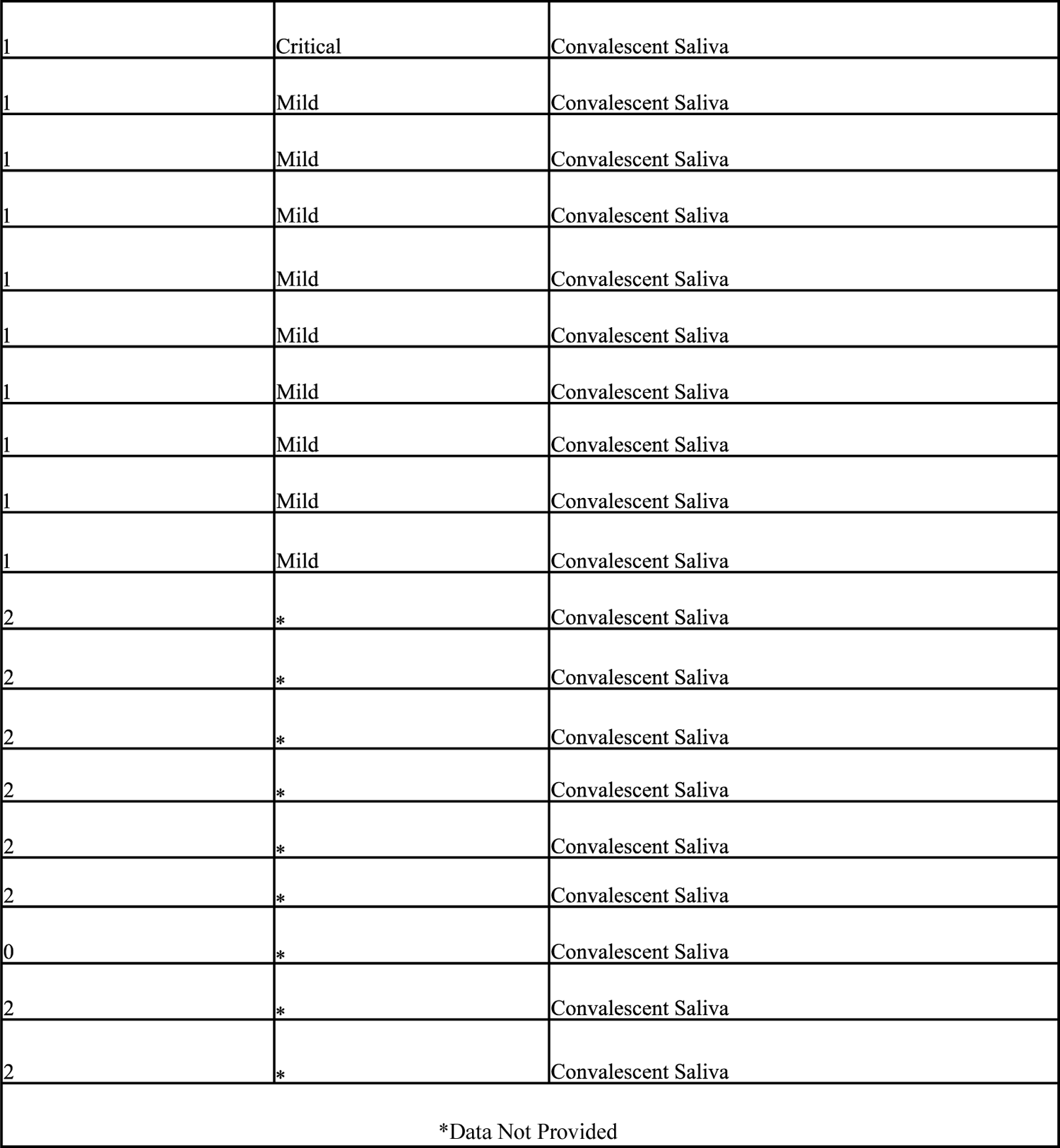
COVID-19 Convalescent Saliva Samples

**Sup. Table 5:**
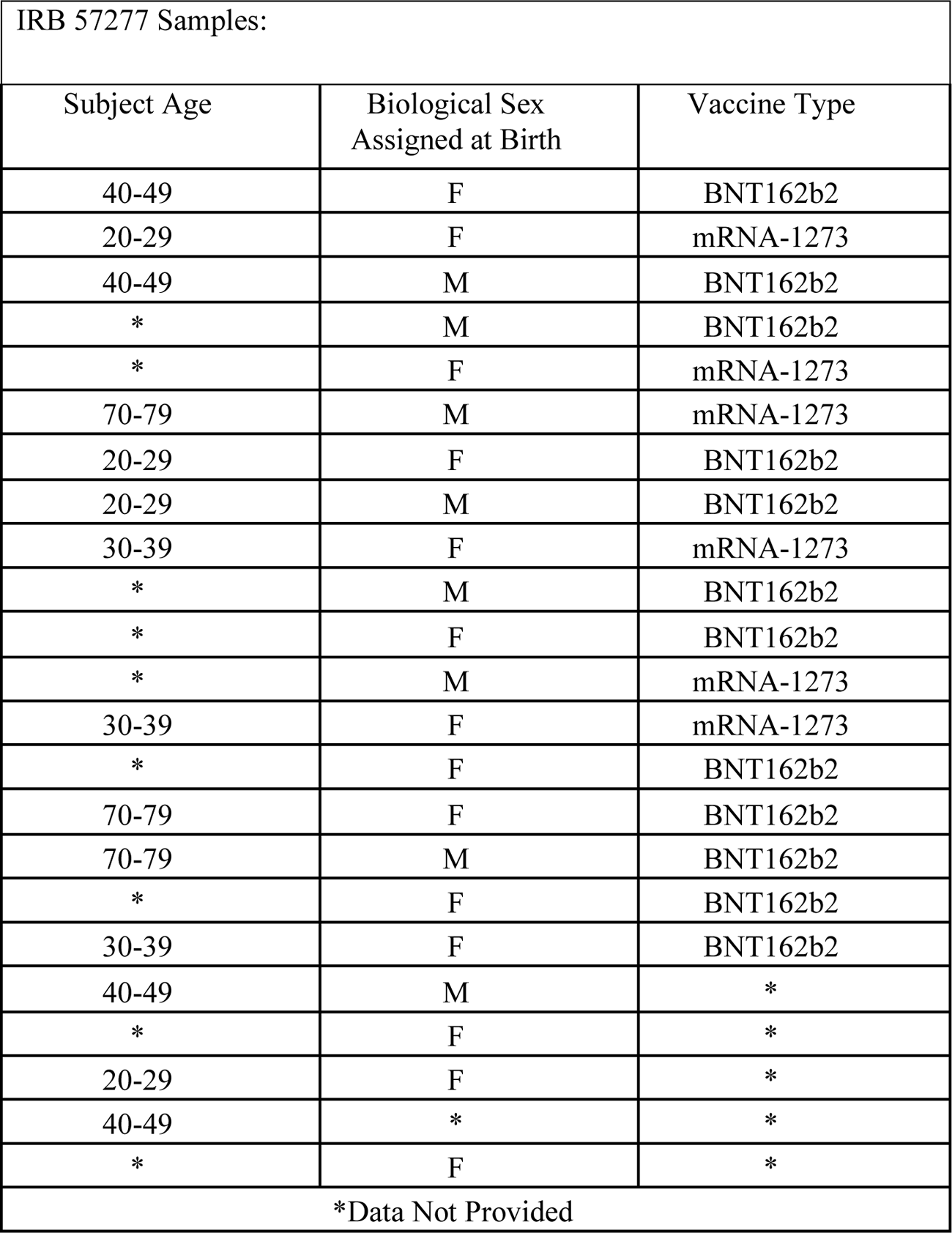
BNT162b2 or mRNA-1273 Saliva

**Sup. Table 6:** Known Negative Saliva Samples

| IRB 57277 Samples: |  |  |
| --- | --- | --- |
| Subject Age | Biological Sex Assigned at Birth | Sample Type |
| 40-49 | F | Known Negative |
| 40-49 | M | Known Negative |
| 70-79 | M | Known Negative |
| 30-39 | F | Known Negative |
| 80-89 | M | Known Negative |
| 30-39 | F | Known Negative |
| 30-39 | F | Known Negative |

